# Age- and Sex-Adjusted Myocardial Flow Reserve Percentiles for Personalized Cardiovascular Risk Assessment

**DOI:** 10.64898/2025.12.30.25343223

**Authors:** Lee Joseph, Ludovic Trinquart, Diana M. Lopez, Simone Brandao, Jenifer M. Brown, Sanjay Divakaran, Daniel Huck, Brittany Weber, Leanne Barrett Goldstein, Jon Hainer, Sylvain Carre, Mark Lemley, Giselle Ramirez, Joanna X. Liang, Ron Blankstein, Sharmila Dorbala, Erick Alexanderson, Isabel Carvajal-Juarez, Wanda Acampa, Rene R. S. Packard, Viet T. Le, Steve Mason, Stacey Knight, Panithaya Chareonthaitawee, Samuel Wopperer, Thomas L. Rosamond, Ronny R. Buechel, Andrew J. Einstein, Mouaz H. Al-Mallah, Leandro Slipczuk, Mark I. Travin, Daniel S. Berman, Damini Dey, Piotr J. Slomka, Marcelo F. Di Carli

## Abstract

**Introduction:** Positron emission tomography (PET) myocardial flow reserve (MFR) is a robust indicator of coronary vascular health and a strong predictor of cardiovascular risk. Clinical guidelines typically use fixed MFR thresholds (e.g., <2.0) to stratify risk, yet this approach overlooks individual variation, particularly by age and sex. We aimed to establish age- and sex-adjusted MFR percentiles and to evaluate their prognostic and predictive performance for cardiovascular risk assessment, in comparison with conventional fixed-threshold MFR approach.

**Methods:** Using data from the REFINE PET registry (24,820 patients from 12 sites), we measured PET MFR and derived age- and sex-adjusted MFR reference percentiles using quantile regression in patients without known coronary artery disease. All patients were categorized into percentile-based quartile groups. The primary outcome for prognostic and prediction analyses was major adverse cardiovascular events (MACE), defined as all-cause mortality, myocardial infarction, or heart-failure hospitalization. Time-to-event associations were evaluated using covariate-adjusted survival models, with cumulative incidence and hazard ratios (HR) estimated at 1 and 5 years in the derivation dataset, an independent but similar validation dataset A, and a high-risk validation dataset B. Predictive performance for MACE was assessed using discrimination, calibration, and reclassification metrics, comparing percentile-based models with models using a fixed MFR threshold (<2.0).

**Results:** Among participants (mean age 66.5 years; median follow-up 3.6 years), age- and sex-adjusted MFR quartile groups were strong independent predictors of MACE, with adjusted HR increasing stepwise across quartile groups at both early and later follow-up. At 1 year, HR (95% CI) comparing the lowest to the highest quartile group were 4.06 (3.41–4.82) in the derivation cohort, 3.31 (2.32–4.71) in validation cohort A, and 2.35 (2.05–2.70) in validation cohort B. At 5 years, the corresponding HR were 2.18 (1.86–2.56), 1.77 (1.31–2.40), and 1.59 (1.36–1.86). Percentile-based models demonstrated consistently higher discrimination, better calibration, and greater net reclassification for MACE at both time points compared with fixed-threshold MFR models. Although 67.2% of patients had preserved MFR (>2.0), cardiovascular risk increased steadily across MFR percentiles even within this range.

**Conclusion:** Age- and sex-adjusted MFR percentiles provide a reliable, clinically actionable measure of vascular health, improving cardiovascular risk stratification by better capturing age- and sex-related heterogeneity in vascular risk. Compared with traditional fixed-threshold approaches, MFR percentiles demonstrate improved predictive performance for cardiovascular risk assessment across diverse patient populations.

**CLINICAL PERSPECTIVE:** *What is new?:*

- In a large, multinational real-world registry of more than 24,000 patients undergoing PET MPI, we derived age- and sex-adjusted myocardial flow reserve (MFR) percentiles, establishing a lifespan-based reference framework with age-specific reference values.
- Age- and sex-adjusted MFR percentiles showed a strong, graded association with 1- and 5-year cardiovascular risk and demonstrated improved prognostic and predictive performance compared with fixed MFR thresholds and models without MFR across independent derivation and validation datasets, including a higher-risk cohort.
- Lower adjusted MFR percentiles were consistently associated with higher risk of major adverse cardiovascular events, including all-cause mortality, myocardial infarction, and heart-failure hospitalization.

*What are the clinical implications?:*

- Age- and sex-adjusted MFR percentiles provide a personalized, dynamic measure of coronary vascular health that improves cardiovascular risk stratification by accounting for age- and sex-related heterogeneity.
- A percentile-based approach may help clinicians better identify patients at elevated cardiovascular risk who may warrant closer monitoring or intensified preventive strategies, while avoiding unnecessary intervention in lower-risk individuals.
- By integrating macrovascular and microvascular dysfunction, age- and sex-adjusted MFR percentiles offer a clinically interpretable marker of vascular aging that may inform future research on aging biomarkers, novel therapeutics and cardiovascular risk assessment across the spectrum of atherosclerotic and microvascular disease.

## INTRODUCTION

Cardiovascular disease remains the leading cause of death worldwide, accounting for approximately one-third of deaths in both men and women (1). Although advancing age is a major determinant of cardiovascular risk, chronological age alone does not completely capture the biological vulnerability of the cardiovascular system (2). Measures of biological age, particularly vascular age, which reflect cumulative structural and functional changes in the arteries, may provide a more precise assessment of cardiovascular risk and a more informative guide for preventive and therapeutic interventions. As populations age, improved characterization of vascular aging and its underlying mechanisms has become increasingly important. Yet, current biological age indicators remain fragmented, weakly predictive of cardiovascular events, and fail to outperform traditional tools such as the Framingham Risk Score. These limitations underscore the need for more reliable and clinically actionable markers of cardiovascular aging (3, 4).

Imaging-based physiologic markers of vascular function offer a compelling opportunity to operationalize the concept of vascular aging. Myocardial flow reserve (MFR) is a physiologic, noninvasive marker of cardiovascular risk, defined as the ratio of myocardial blood flow during peak hyperemia to resting flow (5). Positron emission tomography/computed tomography myocardial perfusion imaging (PET/CT MPI) is considered the gold standard for assessing MFR. Beyond its diagnostic utility, MFR provides important prognostic information across the spectrum of ischemic heart disease, capturing both focal and diffuse atherosclerosis as well as microvascular dysfunction. As such, MFR can serve as a robust marker of vascular aging.

Current clinical guidelines typically rely on fixed MFR thresholds, most commonly MFR < 2.0, to stratify cardiovascular risk (5). Yet MFR shows substantial physiological variability, declining with age and differing by sex, with men generally exhibiting higher values than women. Importantly, fixed MFR thresholds were developed primarily for diagnostic and prognostic classification rather than individualized risk prediction. This one-size-fits-all approach ignores physiological variability, limiting their ability to capture the continuous gradients of vascular aging. Although an MFR below 2.0 is strongly associated with adverse outcomes, values above this threshold are not universally benign, particularly in younger individuals, leading to potential risk misclassification (6, 7). These limitations highlight the need for a more individualized approach to MFR-based risk assessment.

Incorporating age- and sex-adjusted MFR percentiles could offer a more precise measure of coronary vascular health, serving as a dynamic tool for assessing vascular aging, refining risk stratification beyond fixed MFR cutoffs, and evaluating the effects of novel vascular therapies. Leveraging data from a large multicenter registry (8), we sought to address three key questions: (1) What are the age- and sex-adjusted distributions of MFR? (2) How do MFR percentiles relate to the risk of major adverse cardiovascular events among patients undergoing PET MPI? and (3) Do percentile-based MFR measures enhance personalized risk prediction compared with the conventional fixed-threshold MFR approach in PET MPI?

## METHODS

### Study Population

The REFINE PET Registry is a multicenter, international registry comprising clinical, procedural, imaging, and follow-up data from patients undergoing PET/CT MPI for the evaluation of suspected or known coronary artery disease (CAD). The registry systematically collects data on demographics, medical history, stress test findings, imaging details, and outcomes. To ensure consistent reporting across sites, standardized definitions, rigorous data quality procedures, and centralized imaging analysis and quantification are employed. The study design has been previously detailed (8).

The REFINE PET Registry enrolled 27,786 patients who underwent PET/CT MPI between 2006 and 2024 across 12 sites in North and Central America and Europe. All imaging data were centrally reprocessed and quantitatively analyzed in a core laboratory. Of 27,786 enrolled patients, 24,820 met eligibility criteria, including age ≥18 years, pharmacologic stress testing, interpretable myocardial blood flow data, no history of cardiac transplantation, and no missing covariates. To enable independent derivation and validation while preserving the distribution of contributing study sites and calendar years, and minimizing potential site- and calendar year–specific effects, participants were randomly assigned to two datasets in an 80:20 ratio, stratified by site and calendar year: dataset 1 (N = 19,876) and dataset 2 (N = 4,944) (Supplemental Figure 1).

Because population-based reference values for MFR typically require large, asymptomatic community-based cohorts, which are not feasible using PET/CT MPI, we defined a relatively low-risk reference cohort within dataset 1.This derivation cohort included patients without known CAD, without prior cardiac interventions or surgeries, and with resting left ventricular ejection fraction (LVEF) > 40% (N = 12,360). This cohort was used to develop age- and sex-adjusted MFR percentile models using quantile regression.

Within dataset 2, the same exclusion criteria were applied to define an independent validation cohort (validation dataset A, N = 3,070). The remaining patients across datasets 1 and 2, i.e., those with known CAD, prior cardiac interventions or surgeries, or resting LVEF ≤40% comprised a higher-risk validation cohort (validation dataset B, N = 9,390). This cohort was used to provide a transparent, “off-label” assessment of age- and sex-adjusted MFR percentiles in patients at higher baseline cardiovascular risk.

### PET Imaging

All patients underwent PET/CT imaging using standard clinical scanners. Details of imaging protocols have been previously reported (8). Briefly, patients were instructed to abstain from caffeine, methylxanthines, and related medications for at least 12-24 hours prior to the scan. Pharmacologic stress was induced with adenosine, regadenoson, dipyridamole, or dobutamine, selected based on site preference and patient characteristics. Myocardial blood flow (MBF) at rest and during hyperemia was measured using either rubidium-82 or N-13 ammonia as the perfusion radiotracer.

All imaging studies underwent rigorous quality control process at the central core laboratory including manual adjustment of auto-segmented myocardium if needed, verification of PET/CT registration, and automated motion correction of dynamic flow data using commercially available software (Quantitative PET software package [QPET], Cedars-Sinai, Los Angeles) (8–11). PET images were quantified in QPET using batch-processing mode. Imaging variables were extracted from both stress and rest phases across perfusion, gated, and dynamic datasets, and were analyzed globally and, when applicable, by vascular territory, myocardial wall, and segment.

### Study Variables

Maximal hyperemic and resting MBF (expressed in mL·g⁻¹·min⁻¹) were extracted from dynamic PET imaging series using compartmental tracer kinetic modeling. MFR was calculated as the ratio of maximal hyperemic MBF to resting MBF, assessed for the entire left ventricle.

Using a standardized 5-point scoring system applied to 17 myocardial segments, summed rest score (SRS) and summed stress score (SSS) were calculated by summing the quantitative segmental scores from gated myocardial perfusion images acquired at rest and stress, respectively. The difference between these scores was recorded as the summed difference score (SDS). Higher SRS, SSS, and SDS values indicate greater extent of scar, combined ischemia and/or scar, and ischemia, respectively. Each score was then expressed as a percentage of the total myocardium by dividing with the maximum possible score (68) and multiplying by 100%. Resting LVEF was derived from gated perfusion images.

To generate coronary artery calcium (CAC) scores, a validated deep learning model was applied to ungated CT attenuation correction (CTAC) scans for image segmentation and quantification of CAC (12, 13).

Covariates included age, sex, hypertension, diabetes mellitus, hyperlipidemia, smoking status, body mass index, CAC score, SSS, resting LVEF, prior myocardial infarction (MI) or coronary revascularization [included only for validation dataset B], early coronary revascularization, and study site.

### Clinical outcomes

Clinical outcomes were defined using standardized REFINE registry criteria (14). The primary outcome was major adverse cardiovascular events (MACE), defined as a composite of all-cause mortality, non-fatal MI, or hospitalization for heart failure (HF). Secondary endpoints included all-cause mortality, a composite of cardiovascular mortality, non-fatal MI, or HF hospitalization, and cardiovascular mortality.

All-cause and cardiovascular mortality were determined by the study sites using death certificates, the Social Security Death Index or the National Death Index (in the United States), hospital records, or physicians’ documentation. Nonfatal MI was defined as new or worsening chest pain leading to hospitalization with biochemical evidence of myocardial injury confirmed by elevated cardiac enzymes. Hospitalization for HF was identified using the primary discharge diagnosis based on validated International Classification of Diseases (ICD) codes. Early coronary revascularization was defined as percutaneous coronary intervention (PCI) or coronary artery bypass grafting (CABG) performed within 90 days of the PET scan. The time of the initial PET/CT MPI was designated as time zero. We censored follow-up at the time of last patient contact.

### Statistical Analysis

Baseline characteristics were summarized separately for the derivation dataset, validation dataset A, and validation dataset B. Continuous variables are presented as mean ± standard deviation (SD), and categorical variables as counts and percentages.

Age- and sex-adjusted MFR reference percentiles were estimated using quantile regression in patients without known CAD (the derivation dataset) and then applied to all participants. Models included sex, age modeled using restricted cubic splines with 4 knots to allow for nonlinear associations, and an interaction term between sex and spline-transformed age to allow the effect of age on MFR to vary by sex. Predicted marginal estimates and 95% confidence intervals were generated from the 1st through 99th conditional percentiles of MFR across age and sex. Because myocardial flow characteristics differ by radiotracer, MFR was modeled both with and without adjustment for radiotracer type, allowing comparison between age- and sex-adjusted versus age-, sex-, and radiotracer-adjusted MFR percentiles (5).

Every participant was assigned an age- and sex-adjusted MFR percentile using the reference quantile regression model and categorized into one of four quartile groups: ≥ 75th, 50–75th, 25–50th, or < 25th percentile. For comparison with conventional practice, participants were also classified using a fixed MFR threshold (<2.0 vs ≥2.0).

Associations between MFR percentiles and cardiovascular outcomes were evaluated separately within the derivation dataset, validation dataset A, and validation dataset B. Analyses were prespecified for 4 outcomes: (1) MACE; (2) all-cause mortality; (3) a composite of cardiovascular mortality, non-fatal MI, or HF hospitalization; and (4) cardiovascular mortality.

Kaplan–Meier methods were used to estimate event-free survival for MACE and all-cause mortality across MFR quartile groups, with comparisons performed using log-rank tests for trend. For cardiovascular mortality and the composite of cardiovascular mortality, non-fatal MI, or HF hospitalization, cumulative incidence function accounting for the competing risk of non-cardiovascular death were estimated and compared using Gray’s test.

For multivariable modeling, associations were first assessed using Cox proportional hazards models for MACE and all-cause mortality, and Fine–Gray subdistribution hazard models for cardiovascular mortality and the composite of cardiovascular mortality, non-fatal MI, or HF hospitalization. The proportional hazards assumption was evaluated using Schoenfeld residuals and diagnostic plots and was found to be violated. Accordingly, interaction terms between MFR quartiles and log-transformed time were incorporated to model time-varying effects. Mixed-effects Cox or Fine–Gray models were fitted with study site included as a random intercept to account for clustering. All multivariable models adjusted for demographic and clinical covariates, including age, sex, hypertension, diabetes mellitus, hyperlipidemia, smoking status, body mass index, CAC score, SSS, resting LVEF, prior MI or coronary revascularization (included only for validation dataset B), early coronary revascularization, and study site. Hazard ratios (HRs) were estimated at 1- and 5-year time points, with 95% confidence intervals calculated using delta-method approximations.

Predictive performance for MACE was compared across 3 modes: (1) a non-MFR model as the reference, (2) a fixed-threshold MFR model, and (3) an age- and sex-adjusted MFR model. The non-MFR model included clinical and imaging covariates but excluded MFR and study site; the latter was omitted to avoid incorporating non-generalizable, study site-specific effects into risk prediction. The fixed-threshold MFR model added a binary MFR <2 indicator with time-varying effects. The percentile-based model replaced the binary indicator with age- and sex-adjusted MFR quartiles, also modeled with time-varying effects.

Model discrimination was assessed using time-dependent area under the curve (AUC) and concordance index (C-index); overall performance using R², Somers Dxy rank correlation (Dxy), and the Brier score; and calibration by comparing predicted versus observed event probabilities. Net Reclassification Improvement (NRI) was calculated using clinically relevant risk thresholds (∼0.5–5% annualized risk), consistent with prior cardiovascular risk prediction studies (15). Internal validation and calibration were performed using 1,000 bootstrap resamples within each dataset.

Missing data accounted for <3.9% of observations across variables included in multivariable models and we performed complete case analysis. All analyses were conducted using R software (version 4.4.2). All tests were two-sided, and P < 0.05 was considered statistically significant. The study was approved by the institutional review boards of all participating sites. Overall study approval was granted by the Cedars-Sinai Medical Center institutional review board. The study was conducted in accordance with the Declaration of Helsinki, and each site obtained either written informed consent or a waiver of consent for the use of de-identified data.

## RESULTS

### Baseline Characteristics

Among the 24,820 patients from the REFINE PET registry included in this study, the derivation (N = 12,360), validation A (N = 3,070), and validation B (N = 9,390) datasets were similar in mean age (65.3, 65.3, and 68.5 years) and self-reported race (White: 81.2%, 81.1%, and 85.3%). The derivation and validation A datasets demonstrated comparable sex distribution (50.0% and 50.8% male) and overall cardiovascular risk profiles. By design, validation dataset B represented a higher-risk cohort, characterized by a greater proportion of men (71.5%), higher prevalence of hypertension, diabetes, dyslipidemia, and more extensive prior coronary disease (83.8%), including previous MI (43.0%) and history of surgical or coronary revascularization (70.3%).

Mean resting LVEF was normal (defined as ≥ 55%) in the derivation and validation A datasets (∼67%) but low-normal (defined as 50-55%) in validation B (52%). CAC burden was markedly higher in validation B (mean 1,573 vs. 367–370), accompanied by lower global MFR (mean 2.2 vs. 2.6), consistent with a higher burden of both macrovascular and microvascular disease in this high-risk cohort (Table 1, Supplemental Table 1).

**Table 1.**
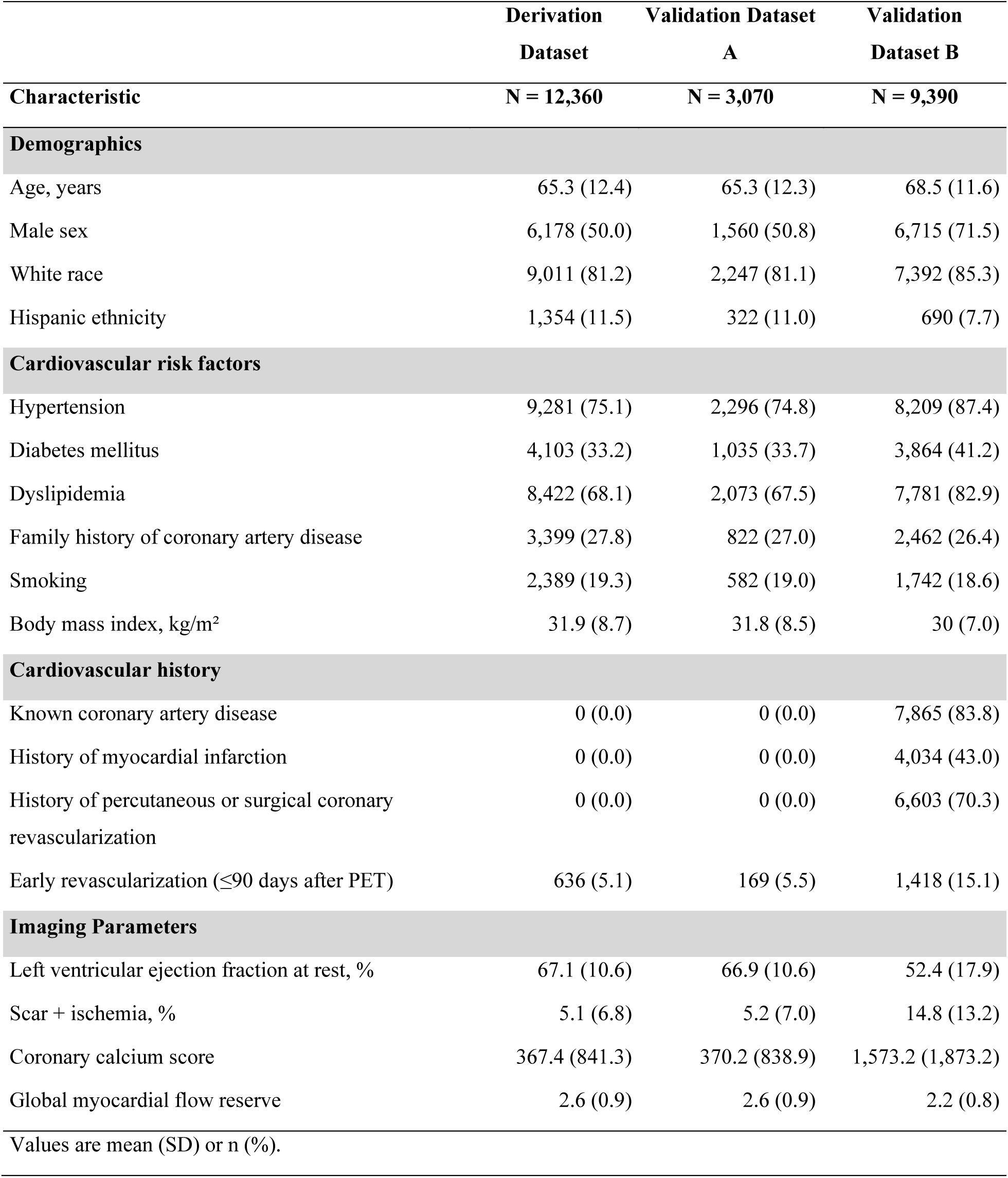
Patient and Imaging Characteristics.

|  | Derivation<br>Dataset | Validation Dataset<br>A | Validation<br>Dataset B |
| --- | --- | --- | --- |
| Characteristic | N = 12,360 | N = 3,070 | N = 9,390 |
| **Demographics** |  |  |  |
| Age, years | 65.3 (12.4) | 65.3 (12.3) | 68.5 (11.6) |
| Male sex | 6,178 (50.0) | 1,560 (50.8) | 6,715 (71.5) |
| White race | 9,011 (81.2) | 2,247 (81.1) | 7,392 (85.3) |
| Hispanic ethnicity | 1,354 (11.5) | 322 (11.0) | 690 (7.7) |
| **Cardiovascular risk factors** |  |  |  |
| Hypertension | 9,281 (75.1) | 2,296 (74.8) | 8,209 (87.4) |
| Diabetes mellitus | 4,103 (33.2) | 1,035 (33.7) | 3,864 (41.2) |
| Dyslipidemia | 8,422 (68.1) | 2,073 (67.5) | 7,781 (82.9) |
| Family history of coronary artery disease | 3,399 (27.8) | 822 (27.0) | 2,462 (26.4) |
| Smoking | 2,389 (19.3) | 582 (19.0) | 1,742 (18.6) |
| Body mass index, kg/m<sup>2</sup> | 31.9 (8.7) | 31.8 (8.5) | 30 (7.0) |
| **Cardiovascular history** |  |  |  |
| Known coronary artery disease | 0 (0.0) | 0 (0.0) | 7,865 (83.8) |
| History of myocardial infarction | 0 (0.0) | 0 (0.0) | 4,034 (43.0) |
| History of percutaneous or surgical coronary revascularization | 0 (0.0) | 0 (0.0) | 6,603 (70.3) |
| Early revascularization ($\leq 90$ days after PET) | 636 (5.1) | 169 (5.5) | 1,418 (15.1) |
| **Imaging Parameters** |  |  |  |
| Left ventricular ejection fraction at rest, % | 67.1 (10.6) | 66.9 (10.6) | 52.4 (17.9) |
| Scar + ischemia, % | 5.1 (6.8) | 5.2 (7.0) | 14.8 (13.2) |
| Coronary calcium score | 367.4 (841.3) | 370.2 (838.9) | 1,573.2 (1,873.2) |
| Global myocardial flow reserve | 2.6 (0.9) | 2.6 (0.9) | 2.2 (0.8) |
| Values are mean (SD) or n (%). |  |  |  |

### MFR Percentile Analysis

Age- and sex-adjusted MFR percentiles demonstrated substantial variation across the lifespan, with systematic shifts in reference values by age and sex (Figure 1). A corresponding table to facilitate individualized clinical interpretation is provided in Supplemental Table 2. Across most ages, men maintained modestly higher MFR values than women, while MFR declined steadily with advancing age in both sexes, with a steeper decrease around age 60 years. Median MFR declined from approximately 3.0 at age 20 years to 2.0 by age 80 years, crossing the conventional abnormal threshold of 2.0 only in the later decades. Adjustment for radiotracer type yielded nearly parallel percentile curves with <5% deviation from age- and sex-adjusted estimates (Supplemental Figures 2-5, Supplemental Table 3), supporting the robustness of age-and sex-adjusted percentiles as a clinically interpretable reference framework without routine radiotracer stratification.

**Figure 1.**
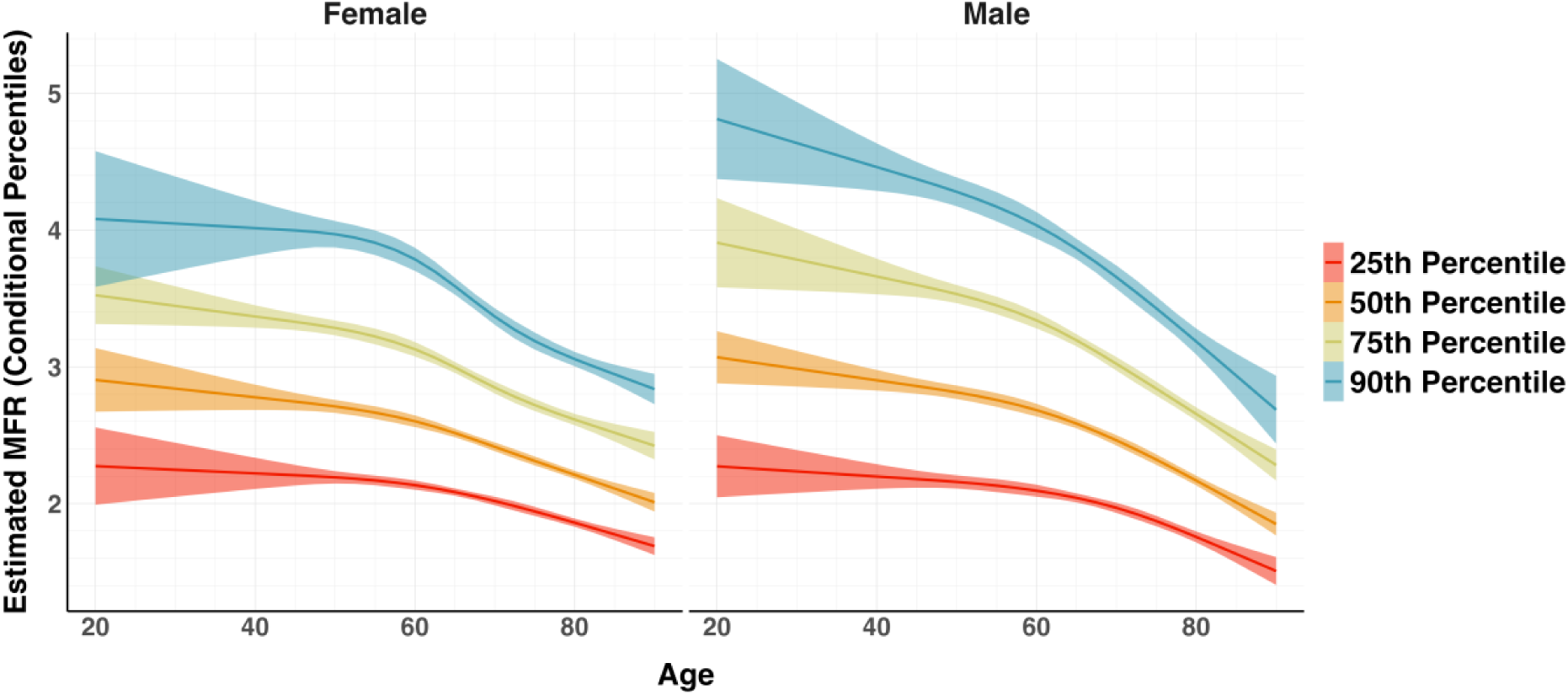
Predicted Age- and Sex-Adjusted MFR Percentiles Estimated Using Quantile Regression. Curves show the adjusted MFR across age, displayed separately for females (left) and males (right). Each line depicts the estimated conditional percentile of MFR adjusted by age and sex. Shaded ribbons show the corresponding 95% confidence intervals.

### Cumulative Incidence by Age- and Sex-Adjusted MFR

Across all datasets, cumulative incidence curves demonstrated clear and graded separation of event incidence across MFR quartiles, which persisted after multivariable adjustment, with higher event rates observed in progressively lower quartile groups (Figures 2 and 3; Supplemental Figures 6 and 7). Log-rank tests for trend were significant for all datasets and outcomes (all P < 0.001). Cumulative incidence functions accounting for competing risks yielded concordant findings, with lower MFR quartile groups associated with higher incidence of a composite of cardiovascular mortality, myocardial infarction, or heart failure hospitalization, and cardiovascular mortality (Figure 4; Supplemental Figures 8 and 9). Together, these findings highlight the consistent and robust prognostic value of age- and sex-adjusted MFR across multiple datasets and clinical outcomes.

**Figure 2.**
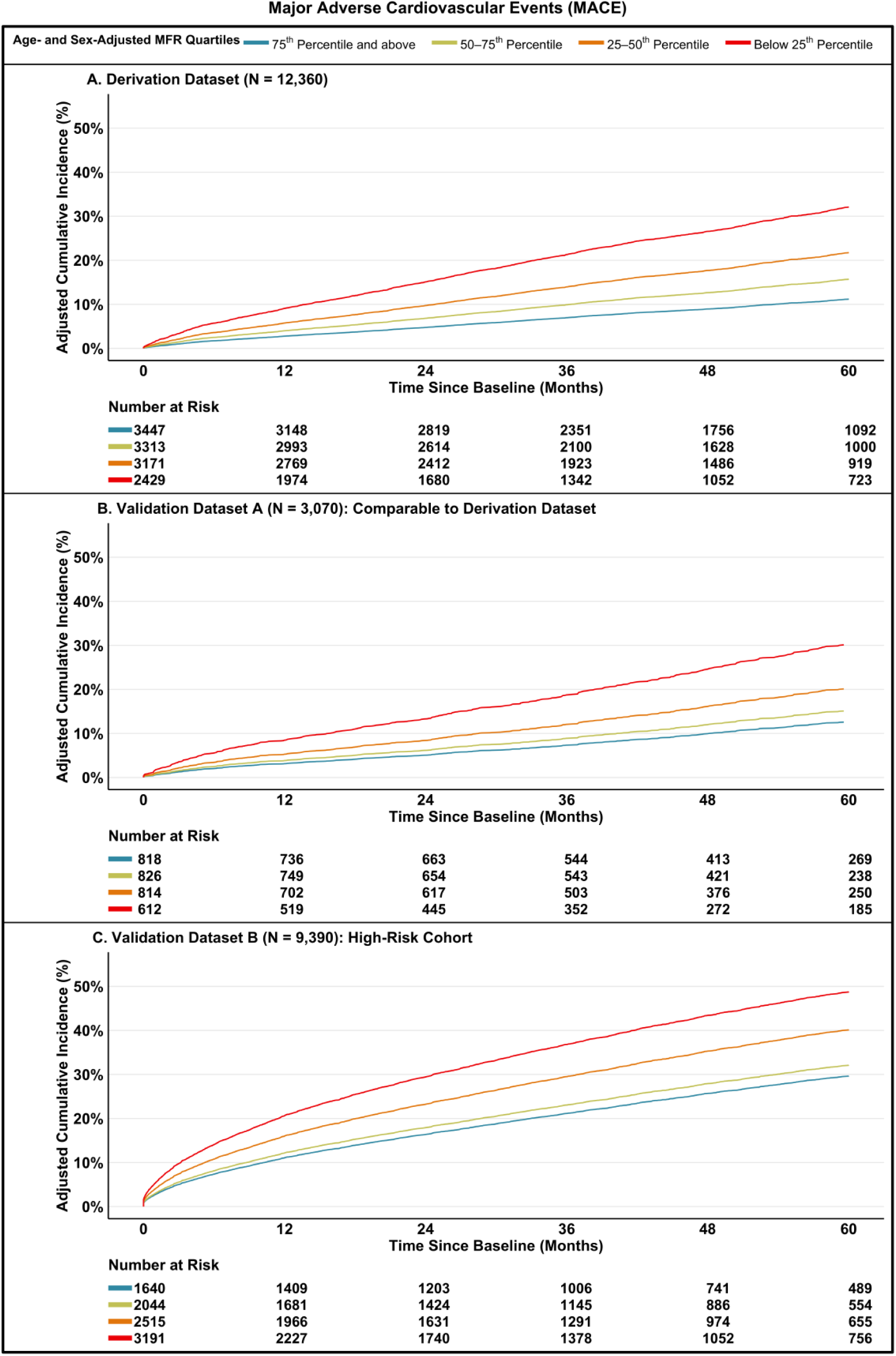
Adjusted Cumulative Incidence of First Major Adverse Cardiovascular Events. Shown is the adjusted 5-year cumulative incidence of first major adverse cardiovascular events among individuals in the derivation (Panel A), validation A (Panel B) and validation B (Panel C) datasets according to quartiles of decreasing levels of age- and sex- adjusted MFR percentiles.

**Figure 3.**
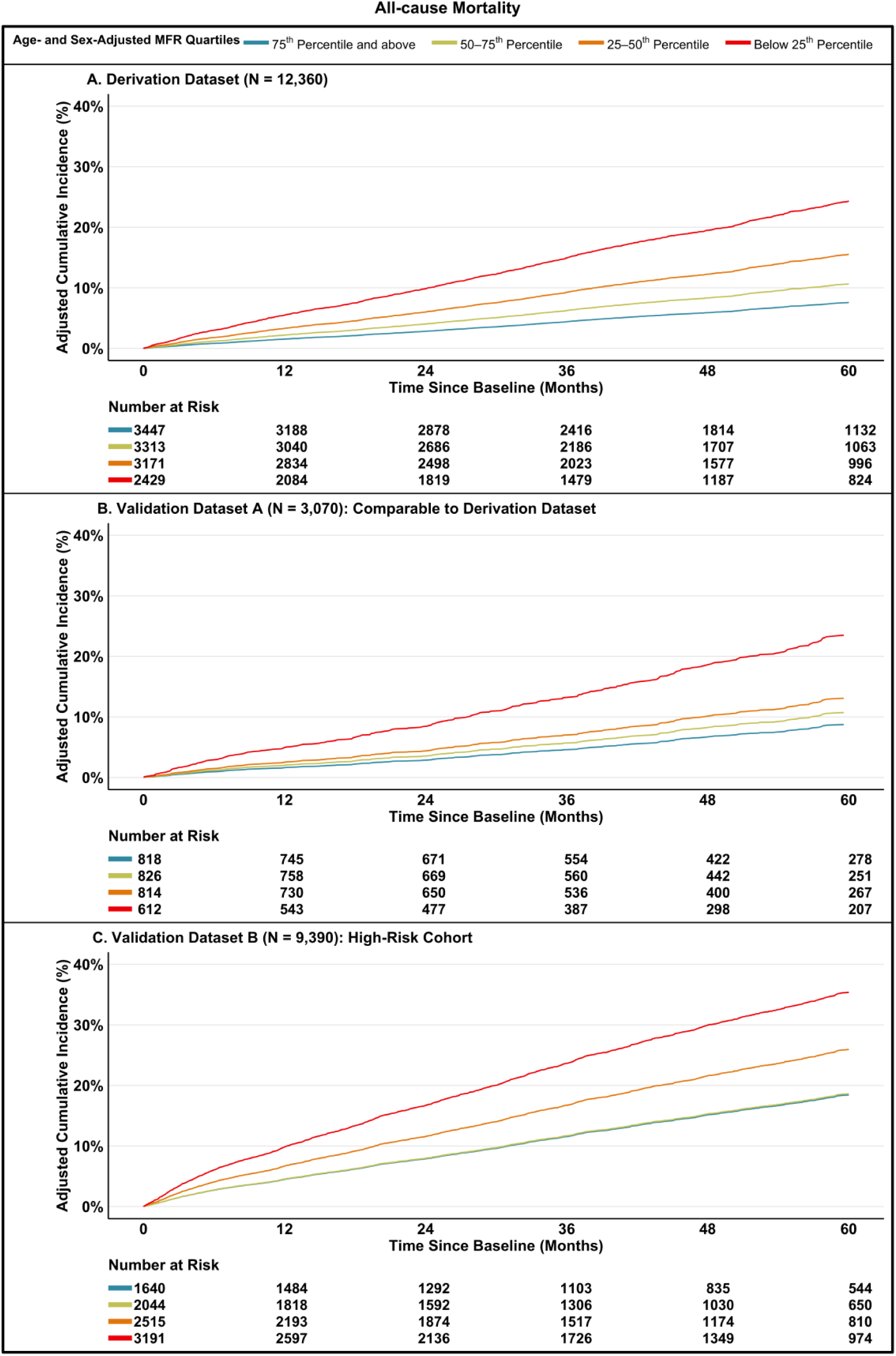
Adjusted Cumulative Incidence of All-cause Mortality. Shown is the adjusted 5-year cumulative incidence of all-cause mortality among individuals in the derivation (Panel A), validation A (Panel B) and validation B (Panel C) datasets according to quartiles of decreasing levels of age- and sex- adjusted MFR percentiles.

**Figure 4.**
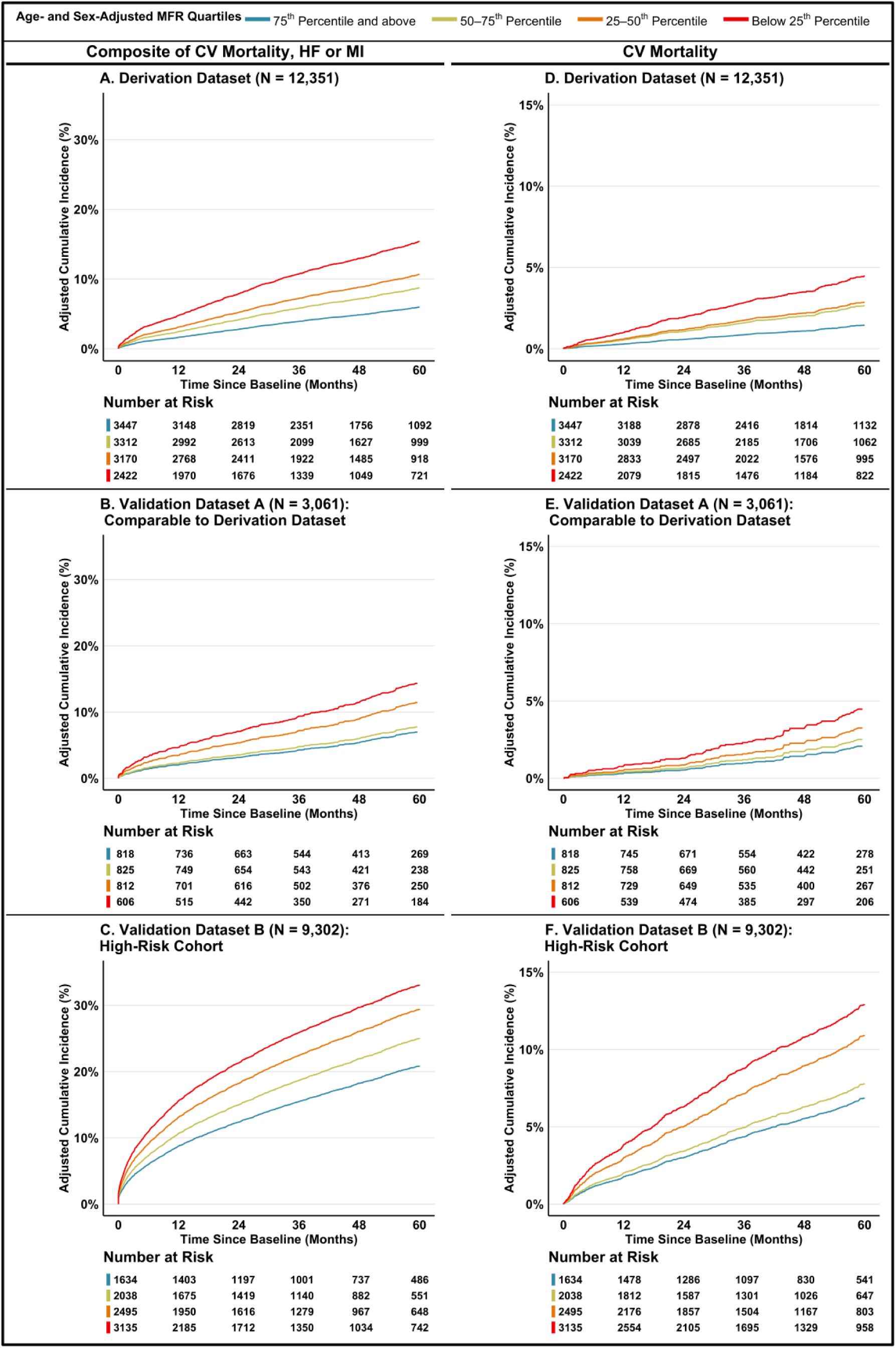
Adjusted Cumulative Incidence of a Composite of Cardiovascular (CV) Mortality, Hospitalization for Heart Failure (HF) or Myocardial Infarction (MI), and CV Mortality. Shown is the adjusted 5-year cumulative incidence of a composite of CV mortality, HF or MI among individuals in the derivation (Panel A), validation A (Panel B) and validation B (Panel C) datasets, and CV Mortality among individuals in the derivation (Panel D), validation A (Panel E) and validation B (Panel F) datasets according to quartiles of decreasing levels of age- and sex-adjusted MFR percentiles.

Throughout each dataset examined, lower age- and sex-adjusted MFR percentile groups had consistently higher incident MACE, all-cause mortality, the composite of cardiovascular mortality, MI, or HF, and cardiovascular mortality (all P < 0.05) (Table 2 and Supplemental Table 4-5). In the derivation dataset, the cumulative incidence of MACE rose stepwise from 2.3% and 10.5% in the top MFR quartile to 12.3% and 33.9% in the lowest quartile at 1 and 5 years, respectively. Similar incremental increases in 1- and 5-year MACE were observed in validation A (2.4% to 9.5%, 11.3% to 33.5%) and validation B (8.1% to 24.2%, 26.5% to 52.9%), paralleling trends for all-cause mortality, cardiovascular mortality, and the composite of cardiovascular mortality, MI, or HF. These findings confirm a robust, graded relationship between lower age- and sex-adjusted MFR and adverse cardiovascular outcomes. Fixed MFR thresholds (<2.0) yielded a broadly similar prognostic signal, while age- and sex-adjusted MFR percentiles provided finer risk resolution and greater discrimination across clinically relevant risk strata (Supplemental Tables 6-8).

**Table 2.** Unadjusted Cumulative Incidence of Major Adverse Cardiovascular Events (MACE) by Age- and Sex-Adjusted MFR Quartile at 1- and 5-Years.

| MACE — Age- and Sex-adjusted MFR Quartile |  |  |  |  |
| --- | --- | --- | --- | --- |
|  | No./Total | 1-Year Cumulative Incidence (% [95% CI]) | 5-Year Cumulative Incidence (% [95% CI]) | P value |
| Derivation Dataset |  |  |  |  |
| ≥75th | 370/3447 | 2.3 (1.8–2.8) | 10.5 (9.2–11.7) | <0.001 |
| 50–75th | 504/3313 | 3.0 (2.4–3.6) | 16.2 (14.6–17.7) |  |
| 25–50th | 691/3171 | 5.5 (4.7–6.4) | 22.5 (20.7–24.3) |  |
| < 25th | 830/2429 | 12.3 (10.9–13.6) | 33.9 (31.7–36.1) |  |
| Validation Dataset A |  |  |  |  |
| ≥75th | 95/818 | 2.4 (1.3–3.5) | 11.3 (8.6–14.0) | <0.001 |
| 50–75th | 125/826 | 3.8 (2.5–5.1) | 15.6 (12.4–18.6) |  |
| 25–50th | 167/814 | 5.9 (4.3–7.6) | 20.8 (17.4–24.0) |  |
| < 25th | 200/612 | 9.5 (7.2–11.9) | 33.5 (29.0–37.8) |  |
| Validation Dataset B |  |  |  |  |
| ≥75th | 409/1640 | 8.1 (6.8–9.4) | 26.5 (23.9–29.0) | <0.001 |
| 50–75th | 594/2044 | 10.6 (9.2–11.9) | 29.8 (27.4–32.1) |  |
| 25–50th | 1013/2515 | 16.3 (14.8–17.7) | 41.4 (39.2–43.7) |  |
| < 25th | 1667/3191 | 24.2 (22.7–25.7) | 52.9 (50.9–54.9) |  |

P-values were obtained by testing for significant differences in time-to-event outcomes across each MFR category, using a log-rank test.

### Association between Age- and Sex-Adjusted MFR and Outcomes

Lower age- and sex-adjusted MFR was strongly and incrementally associated with MACE at both 1 and 5 years across all datasets (Table 3). In the derivation cohort, patients in the lowest quartile had a 4.1-fold higher adjusted risk of 1-year MACE and a 2.2-fold higher risk at 5 years compared with the highest quartile (both P < 0.001). Validation datasets A and B showed consistent patterns, with 1-year adjusted HR of 3.3 in dataset A and 2.4 in dataset B for the lowest versus highest quartile. At 5 years, adjusted HRs were 1.8 in dataset A and 1.6 in dataset B. Intermediate quartiles also exhibited graded risk elevations, underscoring a clear dose–response relationship. Similar graded associations were observed for all-cause mortality, cardiovascular mortality, and the composite of cardiovascular mortality, MI, or HF. These findings confirm that age- and sex-adjusted MFR percentiles provide robust and finely stratified prognostic information for future cardiovascular events.

**Table 3.**
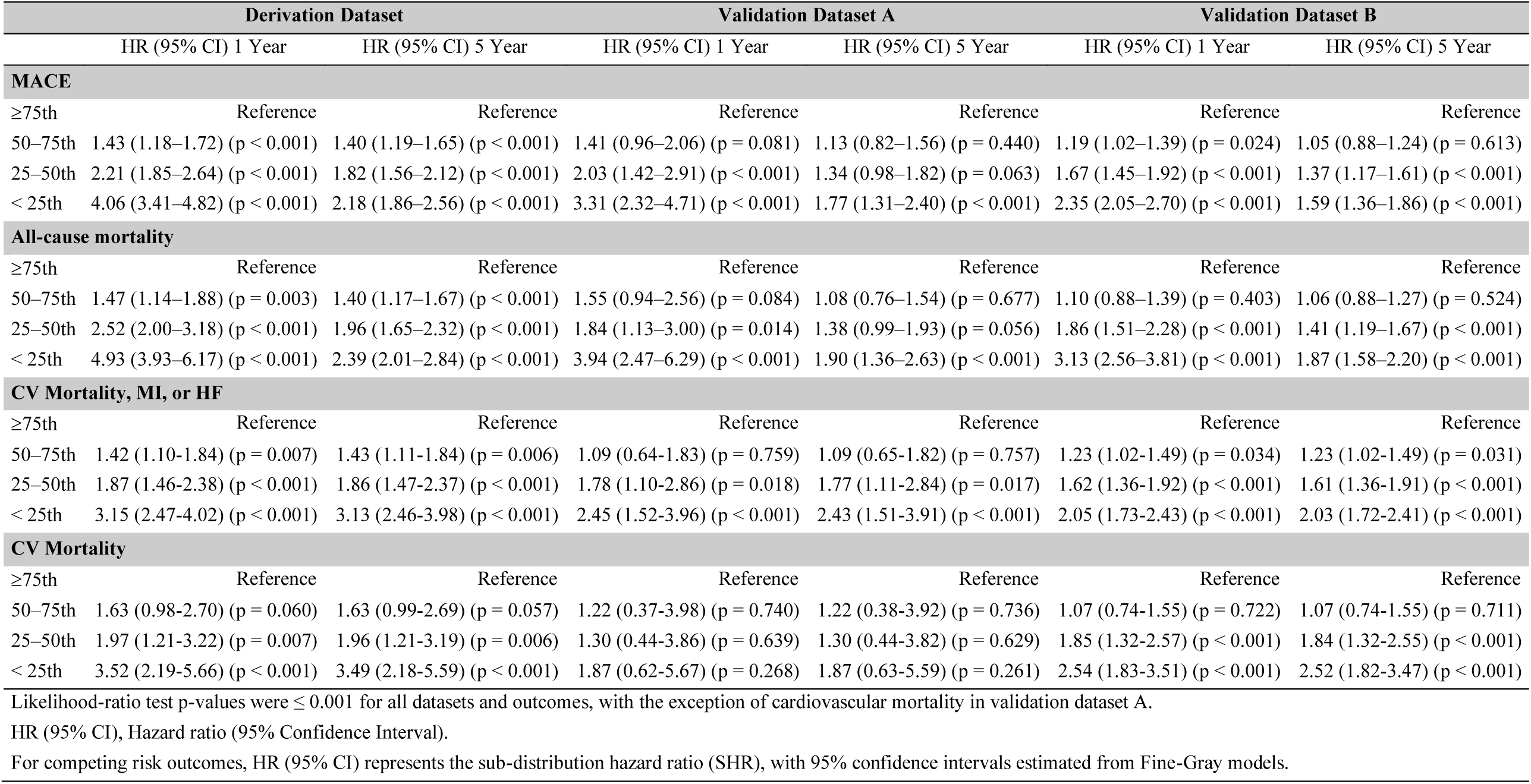
Adjusted Hazard Ratios According to Age- and Sex-Adjusted MFR Quartiles at 1- and 5-Years.

### Model Performance

Across all datasets and at both 1- and 5-year time points, age- and sex-adjusted MFR percentiles consistently and substantially improved risk prediction for MACE compared with models without MFR and those using a fixed threshold (Table 4). Percentile-based models demonstrated higher discrimination, better calibration, and improved overall model performance, outperforming models without MFR and those with a binary MFR threshold. Age- and sex-adjusted MFR percentiles therefore provided more accurate and reliable cardiovascular risk stratification across all analyses, supporting their use as a broadly applicable and clinically meaningful tool for cardiovascular risk prediction.

**Table 4.** Comparative Performance of Models Across Datasets.

|  | Time | Derivation Dataset |  |  | Validation Dataset A |  |  | Validation Dataset B |  |  |
| --- | --- | --- | --- | --- | --- | --- | --- | --- | --- | --- |
|  |  | Non-MFR Model | Fixed MFRThreshold Model | Age- & Sex-Adjusted MFR Model | Non-MFR Model | Fixed MFRThreshold Model | Age- & Sex-Adjusted MFR Model | Non-MFR Model | Fixed MFRThreshold Model | Age- & Sex-Adjusted MFR Model |
| Time-dependent AUC | 1-Year | 0.66 (0.63,0.69) | 0.80 (0.77,0.83) | 0.92 (0.89,0.94) | 0.65 (0.59,0.7) | 0.79 (0.73,0.85) | 0.92 (0.87,0.97) | 0.69 (0.67,0.71) | 0.84 (0.83,0.86) | 0.94 (0.93,0.95) |
|  | 5-Year | 0.68 (0.65,0.69) | 0.82 (0.81,0.84) | 0.94 (0.92,0.95) | 0.68 (0.64,0.73) | 0.84 (0.8,0.87) | 0.94 (0.91,0.97) | 0.70 (0.68,0.72) | 0.90 (0.88,0.91) | 0.96 (0.95,0.97) |
| C-index | 1-Year | 0.66 | 0.80 | 0.91 | 0.65 | 0.78 | 0.91 | 0.67 | 0.82 | 0.93 |
|  | 5-Year | 0.67 | 0.78 | 0.89 | 0.67 | 0.79 | 0.89 | 0.67 | 0.82 | 0.90 |
| Brier Score | 1-Year | 0.05 (0.04,0.05) | 0.03 (0.03,0.03) | 0.01 (0.01,0.01) | 0.05 (0.04,0.06) | 0.03 (0.03,0.04) | 0.01 (0.01,0.02) | 0.13 (0.12,0.13) | 0.07 (0.07,0.08) | 0.02 (0.02,0.03) |
|  | 5-Year | 0.15 (0.14,0.15) | 0.10 (0.10,0.11) | 0.05 (0.04,0.05) | 0.14 (0.13,0.16) | 0.10 (0.09,0.11) | 0.05 (0.04,0.06) | 0.21 (0.21,0.22) | 0.13 (0.12,0.13) | 0.06 (0.05,0.07) |
| NRI | 1-Year | Reference | 0.57 (0.51, 0.62) | 1.11 (1.05, 1.16) | Reference | 0.54 (0.41, 0.66) | 1.10 (0.99, 1.23) | Reference | 0.47 (0.44, 0.49) | 0.75 (0.73, 0.77) |
|  | 5-Year | Reference | 0.29 (0.25, 0.34) | 1.20 (0.99, 1.46) | Reference | 0.33 (0.24, 0.42) | 1.01 (0.74, 1.34) | Reference | 0.23 (0.21, 0.25) | 0.57 (0.5, 0.68) |
| Dxy |  | 0.34 | 0.56 | 0.8 | 0.34 | 0.57 | 0.8 | 0.34 | 0.63 | 0.85 |
| R<sup>2</sup> |  | 0.07 | 0.25 | 0.46 | 0.09 | 0.26 | 0.47 | 0.12 | 0.45 | 0.72 |
| Calibration Slope |  | 0.99 | 1 | 0.99 | 0.96 | 0.98 | 0.97 | 0.99 | 1 | 0.99 |

AUC, Area Under the Curve; C-index, concordance index; NRI, Net Reclassification Improvement; Dxy, Somers' Dxy rank correlation; R<sup>2</sup>, R-squared.
All values are optimism-corrected, with 95% confidence intervals shown in parentheses where applicable.
Metric ranges: 0–1 for AUC, C-index, Brier Score, and R<sup>2</sup>; –2 to +2 for NRI; –1 to +1 for Dxy.

## DISCUSSION

In this large cohort of 24,820 patients undergoing PET/CT MPI, we established age- and sex-adjusted MFR percentiles that define age- and sex-specific reference values across the adult lifespan. These percentiles strongly predicted 1- and 5-year cardiovascular risk, with risk rising across decreasing quartile groups. These findings were consistent in the derivation and the validation datasets. Compared with models without MFR and those using fixed MFR thresholds, percentile-based models demonstrated improved discrimination, calibration, and reclassification across datasets and time horizons, supporting a robust and generalizable framework for cardiovascular risk assessment.

These findings have several implications. First, although traditional models estimate cardiovascular risk, there is growing interest in comprehensive measures that can both assess risk and monitor therapeutic effects across the lifespan (2–4). Extensive evidence supports multiple therapies targeting diverse biological pathways in atherosclerotic disease, including lipid metabolism, vascular inflammation, thrombosis, endothelial function, lipoprotein modification, and glucose and metabolic signaling. In this context, age- and sex-adjusted MFR percentiles integrate multiple biological processes in contrast to traditional single-parameter or score-based approaches and demonstrate strong short- and mid-term predictive performance.

Second, conventional cut-offs, such as MFR <2.0, may overlook intermediate-risk and certain high-risk individuals, whereas percentile-based assessment allows for more nuanced and accurate risk stratification, overcoming a major limitation of fixed-threshold MFR models (6). This is particularly relevant in cardiometabolic and inflammation-driven disease, where identifying subclinical vascular dysfunction could enable earlier use of lipid-lowering or anti-inflammatory therapies to slow vascular aging and support favorable remodeling. Importantly, a single age- and sex-adjusted MFR measurement remained informative for risk estimation up to 5 years, reassuring clinicians concerned about temporal variability.

Third, these percentiles can also serve as a reference marker of cardiovascular risk against which novel aging biomarkers can be validated. They provide a dynamic tool for evaluating anti-aging and vascular-targeted therapies, in contrast to measures with limited reversibility, such as the CAC score. However, additional evidence is needed to clarify how within-individual changes in MFR over time relate to changes in cardiovascular risk, as this represents a potential opportunity for longitudinal risk assessment and treatment monitoring.

As one example, despite the clear benefits of behavioral, pharmacological, and procedural interventions in reducing risk, conditions such as angina with no obstructive CAD (ANOCA) are rising and frequently underrecognized (16, 17). Because ANOCA is not always benign, improved tools for identifying high-risk patients are needed. Age- and sex-adjusted MFR percentiles address this need by identifying patients with relatively low vascular function despite apparently preserved absolute MFR values (e.g., MFR >2.0), facilitating earlier recognition of microvascular dysfunction.

By reclassifying patients into more appropriate risk categories, adjusted percentiles support more precise clinical decision-making and help clinicians identify those who warrant aggressive anti-atherosclerotic therapy. They also provide a quantitative framework for evaluating emerging therapies in future studies, including sodium–glucose cotransporter 2 inhibitors, glucagon-like peptide-1 receptor agonists, and novel ANOCA-targeted therapies. In addition, adjusted percentiles help avoid unnecessary treatment in low-risk individuals, thereby maximizing clinical benefit while minimizing potential harms such as diabetes or pharmacologic hypotension (18–23).

The implications of MFR reference points earlier in life also warrant consideration. Current risk scores typically classify young adults as low risk and would not suggest pharmacologic intervention (24). However, given the rising burden of atherosclerosis in younger populations, adjusted percentiles may still offer valuable insight by confirming that these individuals are indeed at genuinely low risk, particularly in situations of clinical equipoise (25). Younger individuals were underrepresented in this study, and further research is needed to better understand MFR reference points in this population.

This study utilizes a retrospective, multinational cohort with rigorous quality control and centralized processing, combining efficiency in time and cost with high data quality. It includes a large, well-powered sample and addresses a clinically important question with significant implications for cardiovascular risk assessment, providing clinicians and researchers with a valuable tool. Findings are generalizable to patients referred for pharmacologic PET/CT MPI for stable symptoms.

Several limitations should be considered. First, the study population may not represent the broader, non-referral population or specialized groups such as cardiac transplant patients. Second, although missing data was minimal overall, information on abnormal renal function was missing for a substantial proportion of participants and therefore could not be fully adjusted for in the multivariable models. Third, perfusion and flow measurements were fully automatically processed using standard quantitative software, which may differ from semi-automatic measurements, though the increasing adoption of AI-based tools and validated software is likely to standardize automated processing soon. Finally, the proportion of non-White participants was 15–20%, which is lower than that of the overall U.S. population, but may be appropriate given that the study included patients from North and Central America and Europe.

## CONCLUSION

In conclusion, age- and sex-adjusted MFR percentiles provide a robust and clinically interpretable marker of vascular health that strongly predicts 1- and 5-year cardiovascular risk. Compared with models without MFR and fixed-threshold MFR classification, percentile-based approaches demonstrate improved risk stratification and predictive performance, supporting their use for cardiovascular risk assessment and future research on vascular aging.

## Data Availability

All data produced in the present study are available upon reasonable request to the authors.

