## Supplemental File for "Age- and Sex-Adjusted Myocardial Flow Reserve Percentiles for Personalized Cardiovascular Risk Assessment"

[1]Cardiovascular Imaging Program, Brigham and Women's Hospital, Boston, MA 02115, USA
[2]Harvard T.H. Chan School of Public Health, Boston, MA 02115, USA
[3]Tufts Clinical and Translational Science Institute, Tufts University, Boston, MA 02111, USA.
[4] Institute for Clinical Research and Health Policy Studies, Tufts Medical Center, Boston, MA 02111, USA
[5]Harvard Medical School, Boston, MA, 02115, USA
[6]UT Southwestern Medical Center, Dallas, TX 75390, USA
[7]Cedars-Sinai Medical Center, Los Angeles, CA, 90048, USA
[8]National Institute of Cardiology "Ignacio Chavez", Mexico City, Mexico
[9]Federico II University Hospital, Napoli, Italy
[10]David Geffen School of Medicine, University of California, Los Angeles, CA 90095, USA
[11]West Los Angeles Veterans Affairs Medical Center, Los Angeles, CA, 90073, USA
[12]Intermountain Medical Center, Murray, UT 84107, USA
[13]Mayo Clinic, Rochester, MN, 55905, USA
[14]The University of Kansas Health System, Kansas City, KS, 66160, USA
[15]University Hospital Zurich, Zurich, Switzerland
[16]Columbia University/New York-Presbyterian Hospital, New York, 10032, USA
[17]Houston Methodist Academic Institute, Houston, TX, 77030, USA
[18] Montefiore Health System/Albert Einstein College of Medicine, Bronx, NY, 10461, USA
[#]Contributed equally.

SUPPLEMENT

### Supplemental Table 1 – Patient and Imaging Characteristics

| Characteristic | Derivation Dataset N = 12,360 | Validation Dataset A N = 3,070 | Validation Dataset B N = 9,390 |
|---|---|---|---|
| **Medications** | | | |
| β-Blockers | 3,026 (24.5) | 760 (24.8) | 5,022 (53.5) |
| Calcium-channel blockers | 1,863 (15.1) | 478 (15.6) | 1,862 (19.8) |
| Nitrates | 881 (7.1) | 226 (7.4) | 1,858 (19.8) |
| **Indications for PET scan** | | | |
| Preoperative | 1,187 (9.7) | 290 (9.5) | 579 (6.2) |
| Angina | 6,737 (55.0) | 1,651 (54.2) | 4,645 (49.5) |
| Dyspnea | 2,761 (22.5) | 697 (22.9) | 2,125 (22.6) |
| **Serum Laboratories[1]** | | | |
| Estimated glomerular filtration rate (GFR), mL/min/1.73 $m^2$ | 68.8 (27.9) | 69.6 (27.1) | 63.5 (26.2) |
| Low density lipoprotein (LDL) cholesterol, mg/dL | 91.9 (38.4) | 91.5 (36.3) | 77.4 (35.7) |
| High density lipoprotein (HDL) cholesterol, mg/dL | 51.8 (23.9) | 52.7 (24.0) | 45.7 (20.2) |
| **Stress agent** | | | |
| Adenosine | 838 (6.8) | 229 (7.5) | 888 (9.5) |
| Dipyridamole | 396 (3.2) | 93 (3.0) | 320 (3.4) |
| Dobutamine | 106 (0.9) | 25 (0.8) | 76 (0.8) |
| Regadenoson | 11,020 (89.2) | 2,723 (88.7) | 8,106 (86.3) |
| **Radiotracer** | | | |
| Rubidium-82 | 8,409 (68.0) | 2,072 (67.5) | 6,040 (64.3) |
| N-13 ammonia | 3,951 (32.0) | 998 (32.5) | 3,350 (35.7) |
| **Hemodynamic Parameters** | | | |
| Heart rate at rest, bpm | 70.0 (12.7) | 70.5 (12.9) | 69.9 (13.6) |
| Heart rate at peak stress, bpm | 92.9 (17.1) | 93.0 (17.6) | 87.8 (16.9) |
| Mean arterial blood pressure at rest, mm Hg | 92.0 (14.4) | 92.2 (14.7) | 91.6 (15.3) |
| Mean arterial pressure at peak stress, mm Hg | 84.3 (15.4) | 84.5 (15.7) | 82.2 (16.8) |
| Rate pressure product at rest, mm Hg x bpm | 9,484.1 (2,364.9) | 9,537.5 (2,447.3) | 9,348.6 (2,361.6) |
| **Imaging Parameters** | | | |
| Left ventricular ejection fraction at stress, % | 70.1 (11.0) | 70.0 (10.8) | 54.5 (18.0) |
| Scar, % | 2.2 (3.9) | 2.3 (4.1) | 8.4 (10.2) |
| Ischemia, % | 2.9 (4.7) | 3.0 (4.7) | 6.5 (6.9) |
| Global myocardial blood flow at rest, $ml \cdot g^{-1} \cdot min^{-1}$ | 1.0 (0.4) | 1.0 (0.4) | 0.9 (0.3) |

| | | | |
|---|---|---|---|
| Global myocardial blood flow at stress, $ml \cdot g^{-1} \cdot min^{-1}$ | 2.5 (0.8) | 2.5 (0.8) | 1.9 (0.7) |
| **Hospital characteristics** | | | |
| Location | | | |
| Emergency department | 530 (4.3) | 131 (4.3) | 256 (2.8) |
| Inpatient | 2,178 (17.7) | 533 (17.5) | 2,888 (31.7) |
| Outpatient | 9,572 (77.9) | 2,374 (78.1) | 5,954 (65.4) |

Values are mean (SD) or n (%).

[1]Estimated GFR, LDL cholesterol, and HDL cholesterol were missing in 5,476 (44.3%), 5,238 (42.4%), and 4,981 (40.3%) patients in the derivation dataset; 1,356 (44.2%), 1,293 (42.1%), and 1,229 (40.0%) in validation A; and 4,044 (43.1%), 3,685 (39.2%), and 3,560 (37.9%) in validation B, respectively.

Abbreviations: bpm, beats per minute; mm Hg, millimeters of mercury.

**Supplemental Table 2. Age- and Sex-Adjusted Myocardial Flow Reserve Percentiles with 95% Confidence Intervals**

| Percentile | 20 years | 30 years | 40 years | 50 years | 60 years | 70 years | 80 years | 90 years |
|---|---|---|---|---|---|---|---|---|
| **Female** | | | | | | | | |
| 5 | 1.52(1.2,1.84) | 1.53(1.3,1.75) | 1.53(1.4,1.66) | 1.53(1.47,1.59) | 1.52(1.47,1.58) | 1.48(1.44,1.52) | 1.37(1.33,1.41) | 1.22(1.12,1.33) |
| 10 | 1.67(1.46,1.88) | 1.7(1.55,1.84) | 1.73(1.65,1.81) | 1.76(1.72,1.81) | 1.76(1.71,1.8) | 1.68(1.64,1.72) | 1.55(1.52,1.58) | 1.4(1.34,1.47) |
| 15 | 1.89(1.66,2.11) | 1.91(1.75,2.06) | 1.93(1.84,2.02) | 1.94(1.9,1.99) | 1.92(1.88,1.96) | 1.81(1.78,1.84) | 1.66(1.62,1.69) | 1.49(1.41,1.57) |
| 20 | 2(1.78,2.21) | 2.02(1.87,2.17) | 2.05(1.96,2.13) | 2.07(2.02,2.11) | 2.04(2,2.09) | 1.93(1.9,1.97) | 1.78(1.75,1.81) | 1.62(1.54,1.69) |
| 25 | 2.27(1.99,2.56) | 2.25(2.05,2.44) | 2.22(2.11,2.33) | 2.19(2.15,2.24) | 2.13(2.1,2.17) | 2.02(1.99,2.05) | 1.86(1.83,1.89) | 1.69(1.62,1.75) |
| 30 | 2.38(2.2,2.55) | 2.35(2.23,2.47) | 2.32(2.25,2.39) | 2.29(2.25,2.33) | 2.23(2.19,2.26) | 2.11(2.08,2.14) | 1.93(1.9,1.96) | 1.74(1.66,1.81) |
| 35 | 2.5(2.29,2.71) | 2.47(2.32,2.61) | 2.43(2.35,2.52) | 2.4(2.35,2.44) | 2.33(2.29,2.36) | 2.19(2.16,2.22) | 2.01(1.98,2.04) | 1.82(1.74,1.9) |
| 40 | 2.68(2.45,2.91) | 2.62(2.46,2.78) | 2.56(2.47,2.66) | 2.5(2.46,2.55) | 2.41(2.37,2.45) | 2.26(2.23,2.29) | 2.08(2.05,2.11) | 1.9(1.83,1.97) |
| 45 | 2.77(2.55,3) | 2.72(2.56,2.87) | 2.66(2.57,2.75) | 2.6(2.55,2.64) | 2.5(2.46,2.54) | 2.33(2.3,2.36) | 2.15(2.12,2.17) | 1.97(1.91,2.04) |
| 50 | 2.9(2.67,3.14) | 2.84(2.68,3) | 2.78(2.68,2.87) | 2.71(2.66,2.76) | 2.6(2.56,2.64) | 2.41(2.38,2.45) | 2.21(2.18,2.24) | 2.01(1.94,2.08) |
| 55 | 3(2.8,3.2) | 2.94(2.81,3.07) | 2.88(2.8,2.96) | 2.81(2.77,2.86) | 2.7(2.66,2.74) | 2.49(2.46,2.52) | 2.29(2.26,2.33) | 2.11(2.01,2.21) |
| 60 | 3.12(2.9,3.33) | 3.05(2.9,3.2) | 2.98(2.9,3.07) | 2.91(2.87,2.95) | 2.78(2.75,2.82) | 2.56(2.53,2.59) | 2.37(2.33,2.4) | 2.2(2.11,2.29) |
| 65 | 3.27(3.03,3.5) | 3.19(3.02,3.35) | 3.1(3.01,3.2) | 3.02(2.97,3.07) | 2.88(2.83,2.92) | 2.64(2.61,2.68) | 2.45(2.41,2.48) | 2.28(2.2,2.37) |
| 70 | 3.43(3.2,3.67) | 3.34(3.18,3.5) | 3.25(3.16,3.34) | 3.15(3.1,3.21) | 3(2.95,3.05) | 2.74(2.7,2.77) | 2.53(2.5,2.56) | 2.37(2.29,2.44) |
| 75 | 3.52(3.31,3.73) | 3.45(3.3,3.59) | 3.37(3.29,3.45) | 3.28(3.23,3.34) | 3.13(3.08,3.18) | 2.85(2.8,2.89) | 2.61(2.57,2.66) | 2.42(2.32,2.52) |
| 80 | 3.64(3.37,3.91) | 3.58(3.4,3.77) | 3.52(3.41,3.63) | 3.45(3.39,3.52) | 3.3(3.23,3.36) | 2.99(2.94,3.04) | 2.74(2.7,2.78) | 2.55(2.45,2.64) |
| 85 | 3.66(3.35,3.96) | 3.66(3.46,3.87) | 3.67(3.55,3.79) | 3.66(3.59,3.74) | 3.53(3.46,3.59) | 3.16(3.11,3.21) | 2.88(2.83,2.94) | 2.68(2.54,2.82) |
| 90 | 4.08(3.58,4.58) | 4.05(3.7,4.39) | 4.01(3.82,4.21) | 3.97(3.87,4.07) | 3.79(3.7,3.87) | 3.37(3.31,3.43) | 3.06(3,3.11) | 2.84(2.73,2.95) |
| 95 | 4.59(3.82,5.36) | 4.56(4.03,5.09) | 4.53(4.22,4.84) | 4.49(4.34,4.63) | 4.27(4.13,4.4) | 3.76(3.66,3.86) | 3.45(3.36,3.53) | 3.28(3.08,3.47) |
| **Male** | | | | | | | | |
| 5 | 1.58(1.26,1.89) | 1.54(1.33,1.76) | 1.51(1.38,1.64) | 1.48(1.42,1.54) | 1.44(1.39,1.49) | 1.38(1.35,1.42) | 1.25(1.21,1.29) | 1.08(0.97,1.18) |
| 10 | 1.75(1.58,1.93) | 1.73(1.61,1.85) | 1.7(1.63,1.77) | 1.68(1.62,1.73) | 1.65(1.6,1.7) | 1.6(1.55,1.64) | 1.42(1.38,1.46) | 1.19(1.08,1.29) |
| 15 | 1.9(1.7,2.11) | 1.89(1.75,2.03) | 1.88(1.79,1.96) | 1.86(1.81,1.91) | 1.83(1.79,1.88) | 1.75(1.71,1.78) | 1.55(1.51,1.6) | 1.31(1.2,1.43) |
| 20 | 2.02(1.78,2.27) | 2.03(1.86,2.19) | 2.03(1.94,2.13) | 2.03(1.98,2.08) | 1.99(1.94,2.03) | 1.85(1.82,1.89) | 1.65(1.62,1.69) | 1.44(1.34,1.54) |
| 25 | 2.27(2.05,2.5) | 2.24(2.08,2.39) | 2.2(2.11,2.29) | 2.16(2.11,2.21) | 2.09(2.05,2.14) | 1.97(1.93,2.01) | 1.76(1.72,1.8) | 1.51(1.4,1.61) |
| 30 | 2.45(2.17,2.72) | 2.4(2.21,2.59) | 2.35(2.24,2.46) | 2.3(2.25,2.36) | 2.22(2.17,2.27) | 2.06(2.03,2.1) | 1.84(1.8,1.87) | 1.59(1.5,1.68) |
| 35 | 2.63(2.33,2.93) | 2.56(2.35,2.77) | 2.49(2.37,2.61) | 2.42(2.36,2.48) | 2.32(2.28,2.37) | 2.17(2.13,2.21) | 1.93(1.89,1.98) | 1.67(1.56,1.77) |
| 40 | 2.81(2.57,3.05) | 2.73(2.56,2.89) | 2.65(2.55,2.74) | 2.56(2.51,2.62) | 2.45(2.4,2.5) | 2.26(2.22,2.3) | 2.01(1.98,2.05) | 1.75(1.66,1.84) |
| 45 | 2.94(2.74,3.15) | 2.86(2.72,3) | 2.77(2.69,2.85) | 2.69(2.64,2.74) | 2.56(2.51,2.61) | 2.36(2.32,2.4) | 2.09(2.06,2.12) | 1.8(1.72,1.88) |
| 50 | 3.07(2.88,3.26) | 2.99(2.86,3.12) | 2.9(2.83,2.98) | 2.81(2.76,2.86) | 2.68(2.64,2.73) | 2.46(2.42,2.5) | 2.17(2.13,2.2) | 1.85(1.77,1.93) |
| 55 | 3.12(2.85,3.38) | 3.06(2.88,3.24) | 3(2.89,3.1) | 2.93(2.88,2.99) | 2.81(2.76,2.86) | 2.56(2.52,2.6) | 2.25(2.2,2.29) | 1.91(1.8,2.02) |
| 60 | 3.37(3.05,3.69) | 3.27(3.04,3.49) | 3.16(3.03,3.29) | 3.06(2.99,3.12) | 2.9(2.85,2.96) | 2.66(2.62,2.7) | 2.35(2.3,2.4) | 2.02(1.89,2.15) |
| 65 | 3.58(3.24,3.92) | 3.46(3.23,3.7) | 3.35(3.21,3.49) | 3.23(3.16,3.29) | 3.05(3,3.11) | 2.77(2.73,2.82) | 2.45(2.41,2.5) | 2.13(2.02,2.24) |
| 70 | 3.75(3.46,4.03) | 3.63(3.43,3.83) | 3.51(3.39,3.62) | 3.38(3.32,3.44) | 3.2(3.15,3.26) | 2.9(2.86,2.95) | 2.55(2.51,2.59) | 2.18(2.09,2.27) |
| 75 | 3.91(3.58,4.23) | 3.78(3.56,4.01) | 3.66(3.53,3.79) | 3.53(3.47,3.6) | 3.34(3.28,3.4) | 3.02(2.97,3.07) | 2.66(2.61,2.7) | 2.28(2.17,2.4) |
| 80 | 4.22(3.89,4.54) | 4.06(3.84,4.29) | 3.91(3.78,4.04) | 3.75(3.67,3.82) | 3.53(3.46,3.59) | 3.18(3.13,3.23) | 2.78(2.73,2.82) | 2.36(2.24,2.47) |
| 85 | 4.34(3.9,4.77) | 4.22(3.92,4.52) | 4.11(3.93,4.28) | 3.98(3.89,4.08) | 3.77(3.69,3.85) | 3.39(3.31,3.46) | 2.95(2.87,3.02) | 2.51(2.32,2.7) |
| 90 | 4.81(4.37,5.25) | 4.64(4.34,4.94) | 4.46(4.29,4.63) | 4.28(4.17,4.39) | 4.04(3.94,4.13) | 3.66(3.57,3.74) | 3.18(3.09,3.28) | 2.69(2.44,2.93) |
| 95 | 5.14(4.99,5.29) | 5.03(4.94,5.12) | 4.91(4.82,5) | 4.79(4.65,4.93) | 4.56(4.42,4.7) | 4.12(4.01,4.23) | 3.58(3.49,3.67) | 3.02(2.85,3.2) |

**Supplemental Table 3: Radiotracer-Specific Mean Offset in Predicted MFR Across Percentiles**

| | Δ Predicted MFR[1] | |
|---|---|---|
| **Percentile** | **N-13 ammonia** | **Rubidium-82** |
| 0.05 | 0.002 ± 0.001 | -0.001 ± 0.001 |
| 0.10 | 0.004 ± 0.003 | -0.003 ± 0.003 |
| 0.15 | 0.016 ± 0.004 | -0.006 ± 0.004 |
| 0.20 | 0.012 ± 0.011 | -0.01 ± 0.011 |
| 0.25 | 0.017 ± 0.008 | -0.012 ± 0.008 |
| 0.30 | 0.027 ± 0.014 | -0.018 ± 0.014 |
| 0.35 | 0.037 ± 0.007 | -0.019 ± 0.007 |
| 0.40 | 0.039 ± 0.017 | -0.027 ± 0.017 |
| 0.45 | 0.053 ± 0.014 | -0.031 ± 0.014 |
| 0.50 | 0.066 ± 0.017 | -0.042 ± 0.017 |
| 0.55 | 0.073 ± 0.021 | -0.045 ± 0.021 |
| 0.60 | 0.103 ± 0.012 | -0.038 ± 0.012 |
| 0.65 | 0.095 ± 0.031 | -0.062 ± 0.031 |
| 0.70 | 0.12 ± 0.035 | -0.061 ± 0.035 |
| 0.75 | 0.167 ± 0.026 | -0.07 ± 0.026 |
| 0.80 | 0.162 ± 0.046 | -0.105 ± 0.046 |
| 0.85 | 0.231 ± 0.061 | -0.074 ± 0.061 |
| 0.90 | 0.254 ± 0.049 | -0.11 ± 0.049 |
| 0.95 | 0.337 ± 0.109 | -0.149 ± 0.109 |

[1]Values are mean ± standard deviation (SD).

**Supplemental Table 4. Unadjusted Cumulative Incidence by Age- and Sex-Adjusted MFR Quartile at 1- and 5-Years: All-cause Mortality, A Composite of Cardiovascular (CV) Mortality, Myocardial Infarction (MI), or Hospitalization for Heart Failure (HF), and CV Mortality**

| | Derivation Dataset | | | | Validation Dataset A | | | | Validation Dataset B | | | |
|---|---|---|---|---|---|---|---|---|---|---|---|---|
| | No./Total | 1-Year Cumulative Incidence (% [95% CI]) | 5-Year Cumulative Incidence (% [95% CI]) | P value | No./Total | 1-Year Cumulative Incidence (% [95% CI]) | 5-Year Cumulative Incidence (% [95% CI]) | P value | No./Total | 1-Year Cumulative Incidence (% [95% CI]) | 5-Year Cumulative Incidence (% [95% CI]) | P value |
| **All-cause Mortality — Age- and Sex-adjusted MFR Quartile** | | | | | | | | | | | | |
| ≥75th | 260/3447 | 1.1 (0.7–1.4) | 7.3 (6.2–8.4) | <0.001 | 68/818 | 1.3 (0.5–2.0) | 7.2 (5.1–9.4) | <0.001 | 268/1640 | 3.4 (2.5–4.2) | 16.6 (14.4–18.8) | <0.001 |
| 50–75th | 363/3313 | 1.5 (1.1–2.0) | 11.4 (10.1–12.8) | | 92/826 | 2.6 (1.5–3.7) | 11.5 (8.7–14.2) | | 378/2044 | 3.6 (2.8–4.4) | 18.0 (15.9–20.0) | |
| 25–50th | 527/3171 | 3.3 (2.6–3.9) | 15.9 (14.3–17.4) | | 118/814 | 2.3 (1.2–3.3) | 14.1 (11.1–17.0) | | 698/2515 | 6.5 (5.6–7.5) | 27.3 (25.2–29.4) | |
| < 25th | 628/2429 | 7.5 (6.4–8.6) | 25.0 (23.0–27.0) | | 153/612 | 5.4 (3.5–7.2) | 25.1 (20.9–29.0) | | 1250/3191 | 11.8 (10.6–12.9) | 37.9 (36.0–39.8) | |
| **CV Mortality, MI, or HF — Age- and Sex-adjusted MFR Quartile** | | | | | | | | | | | | |
| ≥75th | 188/3447 | 1.4 (1.0–1.8) | 5.3 (4.4–6.2) | <0.001 | 51/818 | 1.9 (0.9–2.8) | 6.1 (4.1–8.1) | <0.001 | 267/1634 | 6.6 (5.3–7.8) | 17.8 (15.6–19.9) | <0.001 |
| 50–75th | 268/3312 | 2.0 (1.6–2.4) | 8.7 (7.8–9.5) | | 59/825 | 1.7 (0.8–2.7) | 7.5 (5.5–9.5) | | 441/2038 | 9.0 (7.8–10.2) | 22.8 (20.7–25.0) | |
| 25–50th | 338/3170 | 3.0 (2.6–3.4) | 11.2 (10.3–12.1) | | 97/812 | 4.0 (3.1–5.0) | 11.8 (9.8–13.8) | | 713/2495 | 13.1 (11.9–14.3) | 30.3 (28.1–32.4) | |
| < 25th | 431/2422 | 6.5 (6.1–6.9) | 17.6 (16.7–18.5) | | 100/606 | 5.6 (4.6–6.5) | 17.7 (15.7–19.8) | | 1133/3135 | 18.8 (17.6–20.0) | 37.2 (35.0–39.3) | |
| **CV Mortality — Age- and Sex-adjusted MFR Quartile** | | | | | | | | | | | | |
| ≥75th | 53/3447 | 0.2 (0.0–0.3) | 1.2 (0.8–1.7) | <0.001 | 13/818 | 0.5 (0.0–1.0) | 1.2 (0.4–2.0) | 0.001 | 95/1634 | 1.6 (1.0–2.2) | 5.4 (4.1–6.7) | <0.001 |
| 50–75th | 94/3312 | 0.5 (0.3–0.6) | 2.9 (2.4–3.3) | | 21/825 | 0.5 (0.0–1.0) | 2.5 (1.7–3.4) | | 156/2038 | 1.5 (0.9–2.1) | 7.3 (5.9–8.6) | |
| 25–50th | 108/3170 | 0.6 (0.5–0.8) | 3.0 (2.5–3.4) | | 33/812 | 0.1 (0.0–0.6) | 3.9 (3.1–4.8) | | 285/2495 | 2.7 (2.1–3.4) | 11.5 (10.2–12.9) | |
| < 25th | 143/2422 | 1.4 (1.2–1.5) | 5.3 (4.9–5.7) | | 35/606 | 1.0 (0.5–1.5) | 5.5 (4.7–6.4) | | 481/3135 | 4.9 (4.2–5.5) | 14.9 (13.6–16.3) | |

P-values were obtained by testing for significant differences in time-to-event outcomes across each MFR category, using a log-rank test for non-competing outcomes or a Gray's test for competing risks outcomes.

### Supplemental Table 5. Unadjusted Cumulative Incidence by Age- and Sex-Adjusted MFR Quartile: HF and MI

| | **Derivation Dataset** | | | | **Validation Dataset A** | | | | **Validation Dataset B** | | | |
|---|---|---|---|---|---|---|---|---|---|---|---|---|
| | No./Total | 1-Year Cumulative Incidence (% [95% CI]) | 5-Year Cumulative Incidence (% [95% CI]) | P value | No./Total | 1-Year Cumulative Incidence (% [95% CI]) | 5-Year Cumulative Incidence (% [95% CI]) | P value | No./Total | 1-Year Cumulative Incidence (% [95% CI]) | 5-Year Cumulative Incidence (% [95% CI]) | P value |
| **MI — Age- and Sex-adjusted MFR Quartile** | | | | | | | | | | | | |
| ≥75th | 73/3447 | 0.5 (0.2–0.7) | 2.1 (1.5–2.6) | <0.001 | 22/818 | 0.4 (0.0–0.8) | 2.7 (1.3–4.2) | 0.012 | 108/1640 | 2.5 (1.7–3.2) | 7.8 (6.3–9.4) | <0.001 |
| 50–75th | 78/3313 | 0.5 (0.2–0.7) | 2.9 (2.4–3.5) | | 23/826 | 0.6 (0.2–1.1) | 3.1 (1.6–4.6) | | 172/2044 | 3.7 (2.9–4.5) | 9.9 (8.3–11.4) | |
| 25–50th | 101/3171 | 0.9 (0.6–1.1) | 3.9 (3.4–4.5) | | 22/814 | 1.1 (0.7–1.6) | 2.5 (1.0–4.0) | | 253/2515 | 5.3 (4.6–6.1) | 12.0 (10.4–13.6) | |
| < 25th | 134/2429 | 2.0 (1.8–2.2) | 6.5 (5.9–7.0) | | 34/612 | 1.2 (0.8–1.6) | 6.5 (5.0–8.0) | | 423/3191 | 7.3 (6.6–8.1) | 16.8 (15.2–18.3) | |
| **HF — Age- and Sex-adjusted MFR Quartile** | | | | | | | | | | | | |
| ≥75th | 86/3447 | 0.8 (0.5–1.1) | 2.7 (2.0–3.3) | <0.001 | 23/818 | 1.1 (0.4–1.9) | 2.9 (1.5–4.3) | <0.001 | 115/1640 | 2.9 (2.1–3.8) | 8.7 (7.1–10.4) | <0.001 |
| 50–75th | 126/3313 | 1.3 (1.0–1.6) | 4.0 (3.4–4.7) | | 24/826 | 0.8 (0.0–1.5) | 3.5 (2.1–4.9) | | 207/2044 | 4.7 (3.9–5.6) | 11.4 (9.7–13.1) | |
| 25–50th | 175/3171 | 1.8 (1.5–2.1) | 6.1 (5.5–6.8) | | 57/814 | 2.8 (2.1–3.5) | 7.7 (6.3–9.1) | | 366/2515 | 6.7 (5.8–7.5) | 16.6 (14.9–18.3) | |
| < 25th | 219/2429 | 3.7 (3.4–4.0) | 10.1 (9.5–10.8) | | 49/612 | 3.9 (3.2–4.7) | 9.9 (8.5–11.3) | | 604/3191 | 10.0 (9.2–10.9) | 23.0 (21.4–24.7) | |

P-values were obtained by testing for significant differences in time-to-event outcomes across MFR categories, using a Gray's test for competing risks outcomes.

**Supplemental Table 6. Unadjusted Cumulative Incidence by Fixed MFR Threshold: Major Adverse Cardiovascular Events (MACE) and All-cause Mortality**

| | Derivation Dataset | | | | Validation Dataset A | | | | Validation Dataset B | | | |
|---|---|---|---|---|---|---|---|---|---|---|---|---|
| | No./Total | 1-Year Cumulative Incidence (% [95% CI]) | 5-Year Cumulative Incidence (% [95% CI]) | P value | No./Total | 1-Year Cumulative Incidence (% [95% CI]) | 5-Year Cumulative Incidence (% [95% CI]) | P value | No./Total | 1-Year Cumulative Incidence (% [95% CI]) | 5-Year Cumulative Incidence (% [95% CI]) | P value |
| **MACE — Fixed MFR Threshold** | | | | | | | | | | | | |
| Preserved MFR (≥ 2.0) | 1321/9234 | 3.2 (2.8–3.6) | 14.6 (13.8–15.5) | <0.001 | 313/2261 | 3.4 (2.6–4.2) | 13.7 (11.9–15.4) | <0.001 | 1506/5190 | 10.5 (9.7–11.4) | 30.0 (28.5–31.4) | <0.001 |
| Impaired MFR (< 2.0) | 1074/3126 | 11.4 (10.3–12.6) | 34.6 (32.7–36.6) | | 274/809 | 10.0 (7.9–12.0) | 35.3 (31.3–39.1) | | 2177/4200 | 23.4 (22.1–24.7) | 52.9 (51.2–54.6) | |
| **All-cause Mortality — Fixed MFR Threshold** | | | | | | | | | | | | |
| Preserved MFR (≥ 2.0) | 942/9234 | 1.7 (1.4–1.9) | 10.0 (9.3–10.8) | <0.001 | 218/2261 | 1.7 (1.2–2.3) | 9.0 (7.6–10.5) | <0.001 | 938/5190 | 3.5 (3.0–4.0) | 17.4 (16.2–18.7) | <0.001 |
| Impaired MFR (< 2.0) | 836/3126 | 7.0 (6.1–7.9) | 26.0 (24.1–27.8) | | 213/809 | 5.4 (3.8–7.0) | 26.9 (23.1–30.5) | | 1656/4200 | 11.6 (10.6–12.6) | 38.9 (37.2–40.6) | |

P-values were obtained by testing for significant differences in time-to-event outcomes across MFR categories, using a log-rank test.

### Supplemental Table 7. Unadjusted Cumulative Incidence by Fixed MFR Threshold: A Composite of CV Mortality, MI, or HF, and CV Mortality

| | Derivation Dataset | | | | Validation Dataset A | | | | Validation Dataset B | | | |
|---|---|---|---|---|---|---|---|---|---|---|---|---|
| | No./Total | 1-Year Cumulative Incidence (% [95% CI]) | 5-Year Cumulative Incidence (% [95% CI]) | P value | No./Total | 1-Year Cumulative Incidence (% [95% CI]) | 5-Year Cumulative Incidence (% [95% CI]) | P value | No./Total | 1-Year Cumulative Incidence (% [95% CI]) | 5-Year Cumulative Incidence (% [95% CI]) | P value |
| **CV Mortality, MI, or HF — Fixed MFR Threshold** | | | | | | | | | | | | |
| Preserved MFR (≥ 2.0) | 675/9233 | 1.9 (1.7–2.2) | 7.6 (6.9–8.2) | <0.001 | 165/2260 | 2.2 (1.6–2.8) | 7.3 (6.0–8.6) | <0.001 | 1088/5173 | 8.9 (8.1–9.7) | 22.3 (21.0–23.6) | <0.001 |
| Impaired MFR (< 2.0) | 550/3118 | 6.1 (5.8–6.4) | 17.7 (17.1–18.4) | | 142/801 | 5.7 (5.1–6.4) | 18.8 (17.6–20.1) | | 1466/4129 | 18.0 (17.2–18.8) | 36.9 (35.6–38.2) | |
| **CV Mortality — Fixed MFR Threshold** | | | | | | | | | | | | |
| Preserved MFR (≥ 2.0) | 202/9233 | 0.4 (0.3–0.5) | 2.0 (1.6–2.3) | <0.001 | 50/2260 | 0.4 (0.1–0.7) | 1.8 (1.1–2.5) | <0.001 | 364/5173 | 1.5 (1.2–1.8) | 6.4 (5.7–7.2) | <0.001 |
| Impaired MFR (< 2.0) | 196/3118 | 1.2 (1.1–1.4) | 5.7 (5.3–6.0) | | 52/801 | 0.8 (0.5–1.0) | 6.8 (6.1–7.5) | | 653/4129 | 4.8 (4.5–5.2) | 16.0 (15.2–16.7) | |

P-values were obtained by testing for significant differences in time-to-event outcomes across MFR categories, using a Gray's test for competing risks outcomes.

### Supplemental Table 8. Unadjusted Cumulative Incidence by Fixed MFR Threshold: HF and MI

| | Derivation Dataset | | | | Validation Dataset A | | | | Validation Dataset B | | | |
|---|---|---|---|---|---|---|---|---|---|---|---|---|
| | No./Total | 1-Year Cumulative Incidence (% [95% CI]) | 5-Year Cumulative Incidence (% [95% CI]) | P value | No./Total | 1-Year Cumulative Incidence (% [95% CI]) | 5-Year Cumulative Incidence (% [95% CI]) | P value | No./Total | 1-Year Cumulative Incidence (% [95% CI]) | 5-Year Cumulative Incidence (% [95% CI]) | P value |
| **MI — Fixed MFR Threshold** | | | | | | | | | | | | |
| Preserved MFR (≥ 2.0) | 221/9234 | 0.5 (0.4–0.7) | 2.7 (2.3–3.1) | <0.001 | 55/2261 | 0.6 (0.3–1.0) | 2.5 (1.7–3.3) | <0.001 | 444/5190 | 3.9 (3.3–4.4) | 9.9 (8.9–10.9) | <0.001 |
| Impaired MFR (< 2.0) | 165/3126 | 1.9 (1.7–2.0) | 6.3 (5.9–6.7) | | 46/809 | 1.3 (1.0–1.6) | 6.4 (5.6–7.2) | | 512/4200 | 6.7 (6.2–7.3) | 15.6 (14.6–16.6) | |
| **HF — Fixed MFR Threshold** | | | | | | | | | | | | |
| Preserved MFR (≥ 2.0) | 327/9234 | 1.2 (0.9–1.4) | 3.8 (3.3–4.3) | <0.001 | 80/2261 | 1.3 (0.8–1.8) | 3.9 (2.9–4.9) | <0.001 | 517/5190 | 4.4 (3.8–4.9) | 11.3 (10.3–12.3) | <0.001 |
| Impaired MFR (< 2.0) | 279/3126 | 3.5 (3.3–3.7) | 10.1 (9.6–10.5) | | 73/809 | 4.1 (3.7–4.6) | 10.9 (9.9–11.9) | | 775/4200 | 9.7 (9.1–10.2) | 22.6 (21.6–23.6) | |

P-values were obtained by testing for significant differences in time-to-event outcomes across MFR categories, using a Gray's test for competing risks outcomes.

Supplemental Figure 1. Study Flowchart for the REFINE PET Registry Analysis

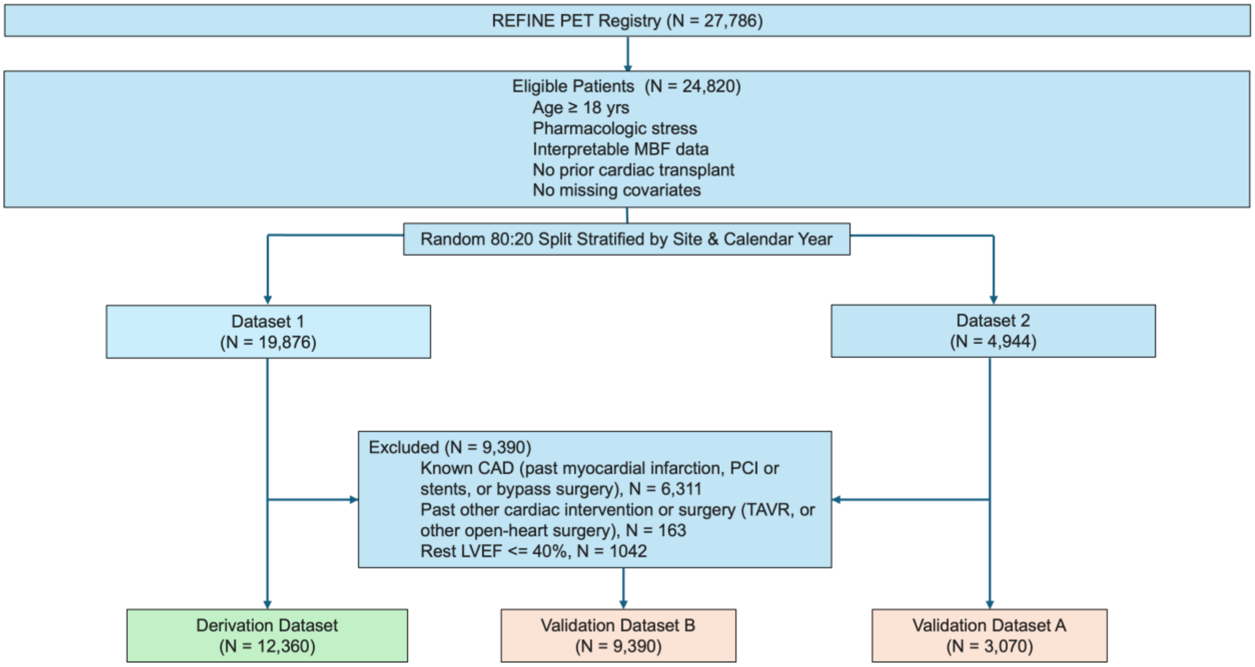


Abbreviations: MBF, myocardial blood flow; CAD, coronary artery disease; PCI, percutaneous coronary intervention; TAVR, transcatheter aortic valve replacement; LVEF, left ventricular ejection fraction.

Supplemental Figure 2. Predicted Age-, Sex- and Radiotracer-adjusted MFR Percentiles Estimated Using Quantile Regression. Curves show the adjusted MFR across age, displayed separately for females (top) and males (bottom), and by radiotracers, N-13 ammonia (left) and Rubudium-82 (right). Each line depicts the estimated conditional percentile of MFR adjusted by age, sex and radiotracer. Shaded ribbons show the corresponding 95% confidence intervals.

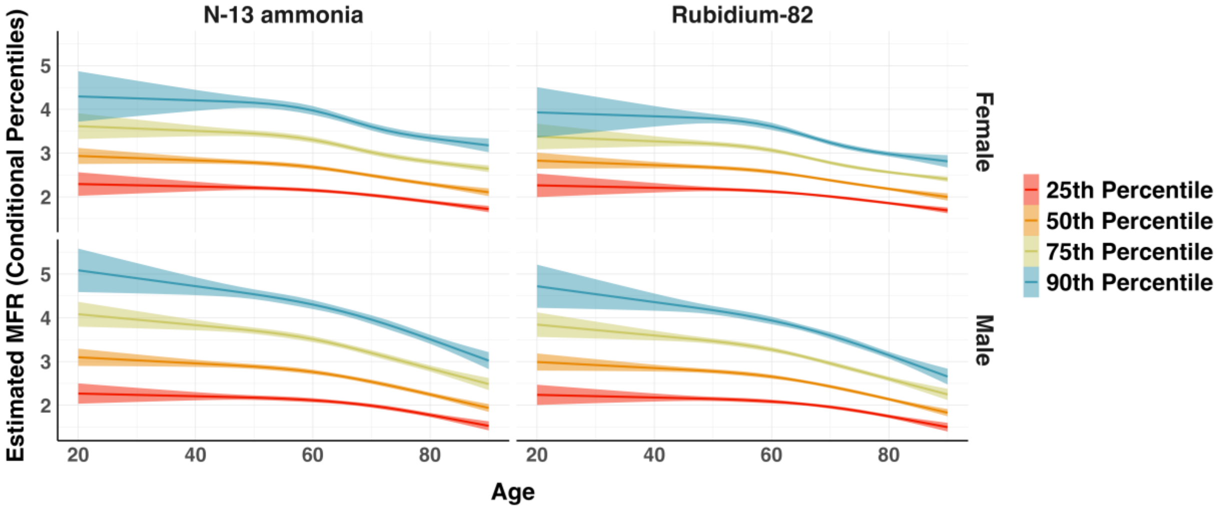

Supplemental Figure 3. Predicted Age-, Sex- and Radiotracer-adjusted MFR Percentiles Estimated Using Quantile Regression. Curves show the adjusted MFR across age, displayed separately for females (left) and males (right), and by radiotracers, N-13 ammonia (blue) and Rubudium-82 (red). Each line depicts the estimated conditional percentile of MFR adjusted by age, sex and radiotracer.

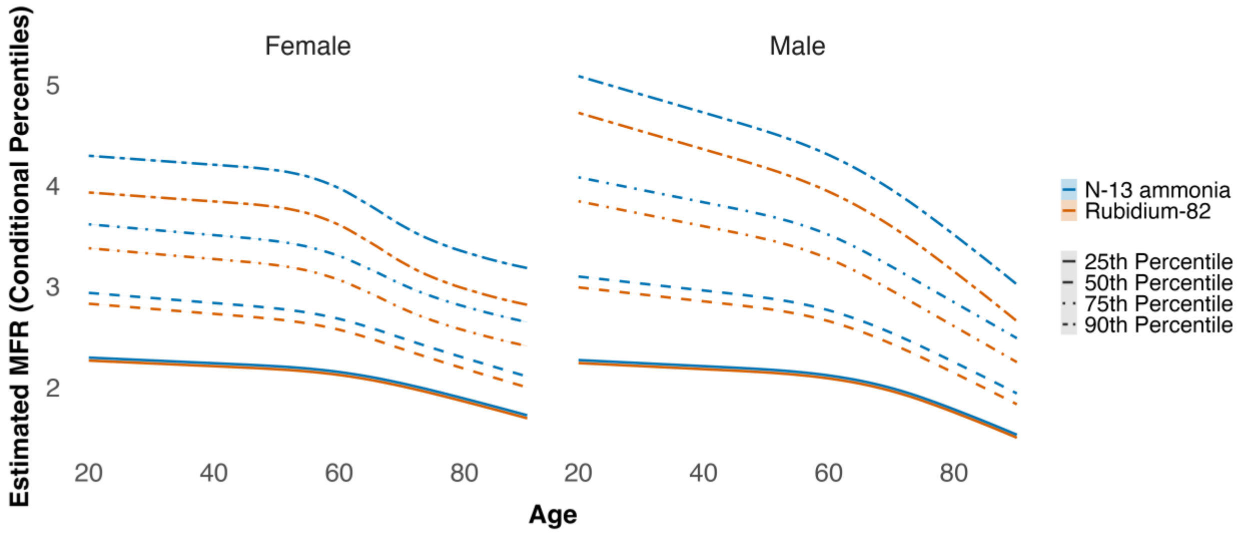

Supplemental Figure 4. Difference in Predicted MFR Between N-13 ammonia and Rubidium-82. Curves show difference in predicted MFR averaged over age and sex, plotted across the full MFR percentile range (1st–99th). The dashed horizontal gray line at 0 denotes no difference between radiotracers. The x-axis represents the percentile of predicted MFR, and the y-axis represents the difference in predicted MFR (N-13 ammonia − Rubidium-82).

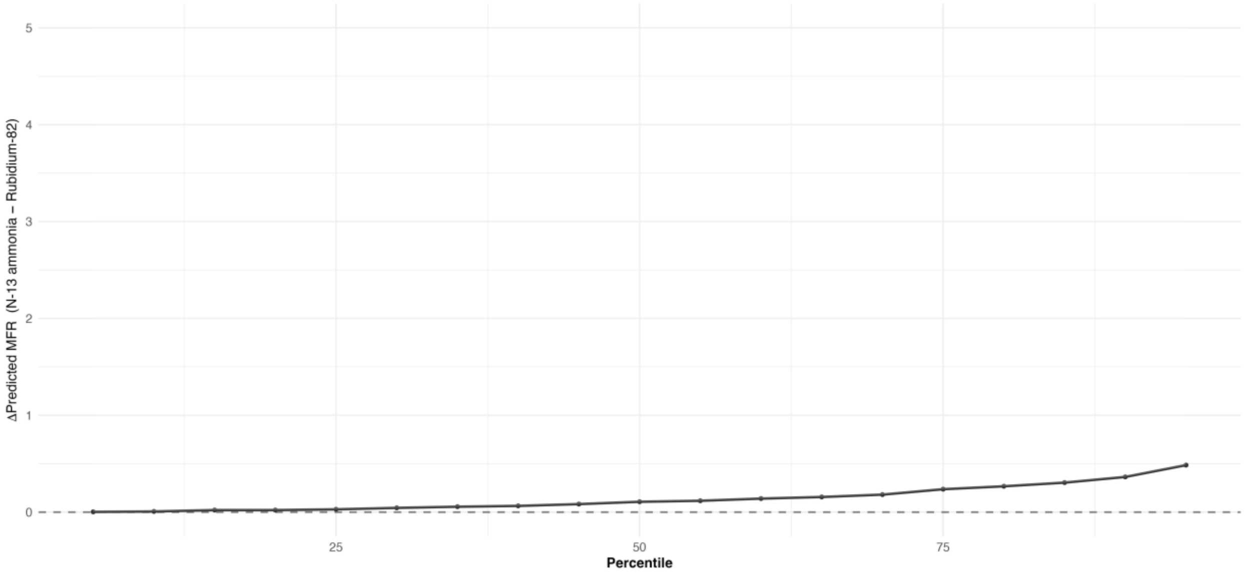

Supplemental Figure 5. Offset in Predicted MFR by Radiotracer Across Percentiles. Curves depict the difference in predicted MFR between the radiotracer-adjusted and radiotracer-unadjusted models, averaged over age and sex, plotted across the full MFR percentile range. The blue curve represents N-13 ammonia, and the red curve represents Rubidium-82. The dashed horizontal gray line at 0 denotes no offset. The x-axis represents the MFR percentile, and the y-axis represents the change in predicted MFR.

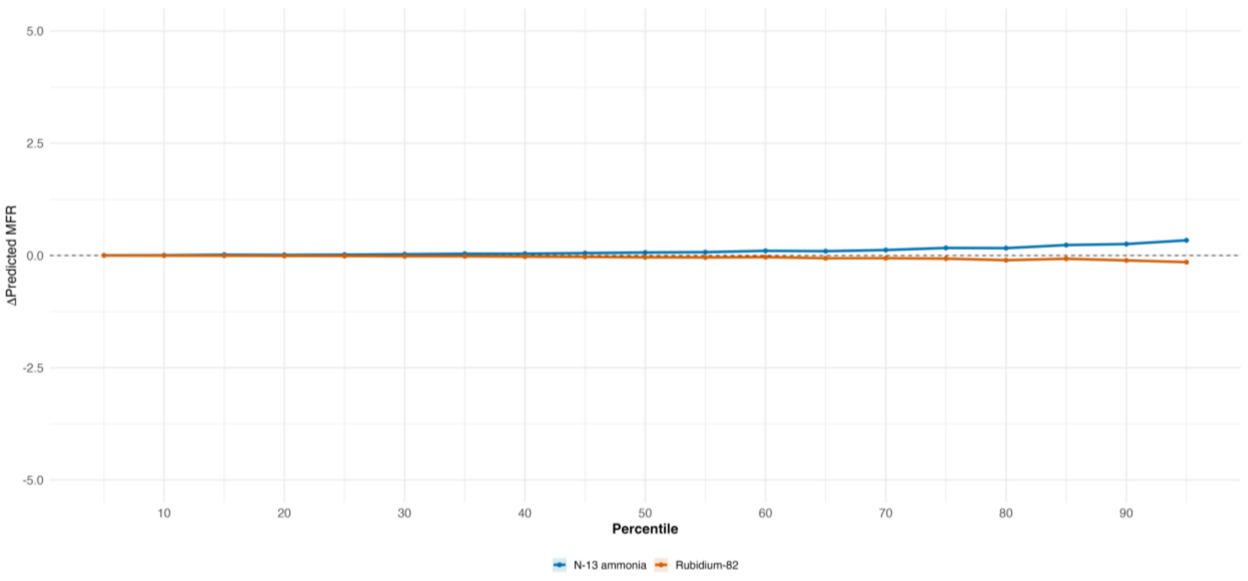

Supplemental Figure 6. Unadjusted Cumulative Incidence of First Major Adverse Cardiovascular Events. Shown is the unadjusted 5-year cumulative incidence of first major adverse cardiovascular events among individuals in the derivation (Panel A), validation A (Panel B) and validation B (Panel C) datasets according to quartiles of decreasing levels of age- and sex-adjusted MFR percentiles.

Major Adverse Cardiovascular Events (MACE)

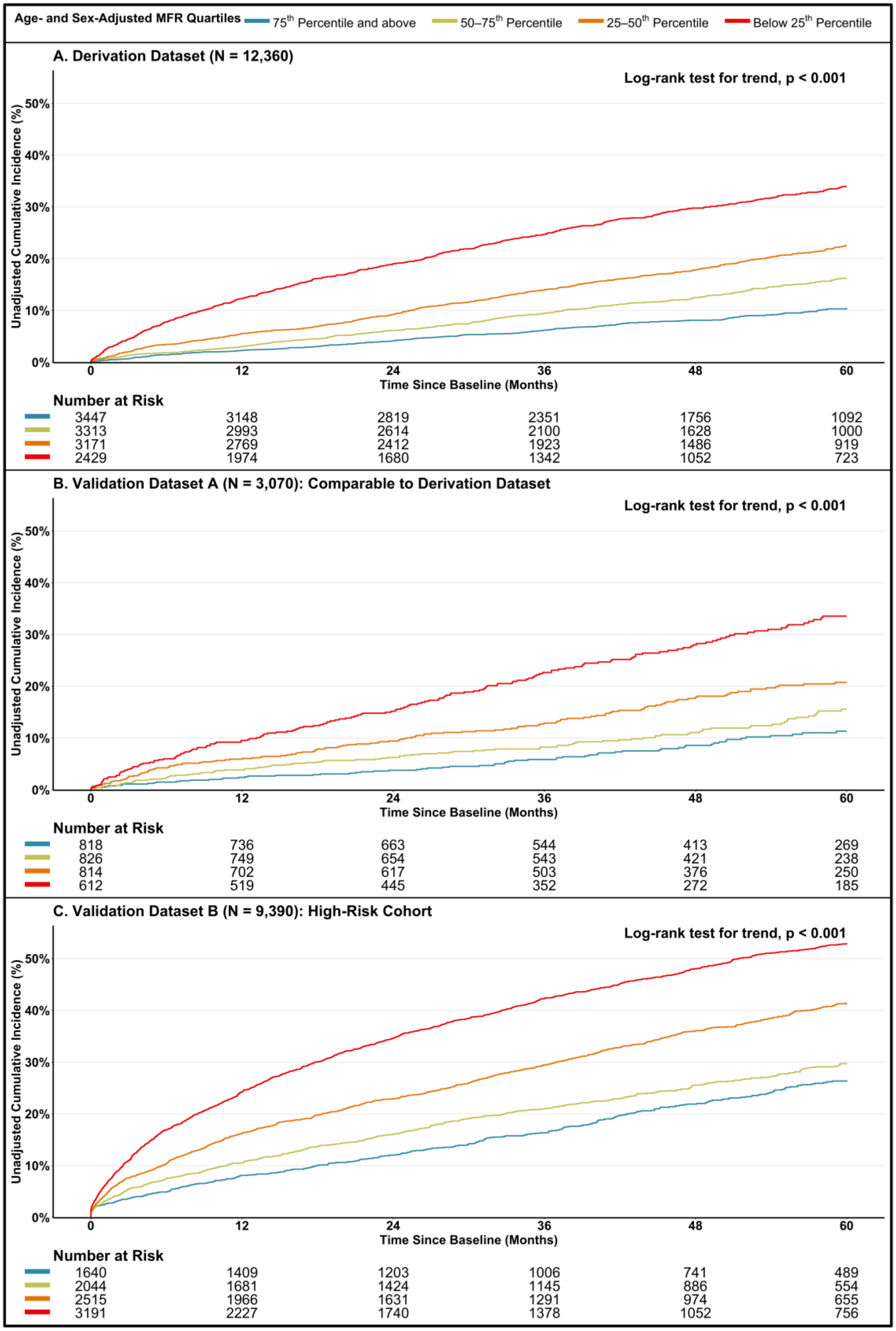

Supplemental Figure 7. Unadjusted Cumulative Incidence of All-cause Mortality. Shown is the unadjusted 5-year cumulative incidence of all-cause mortality among individuals in the derivation (Panel A), validation A (Panel B) and validation B (Panel C) datasets according to quartiles of decreasing levels of age- and sex- adjusted MFR percentiles.

**All-cause Mortality**

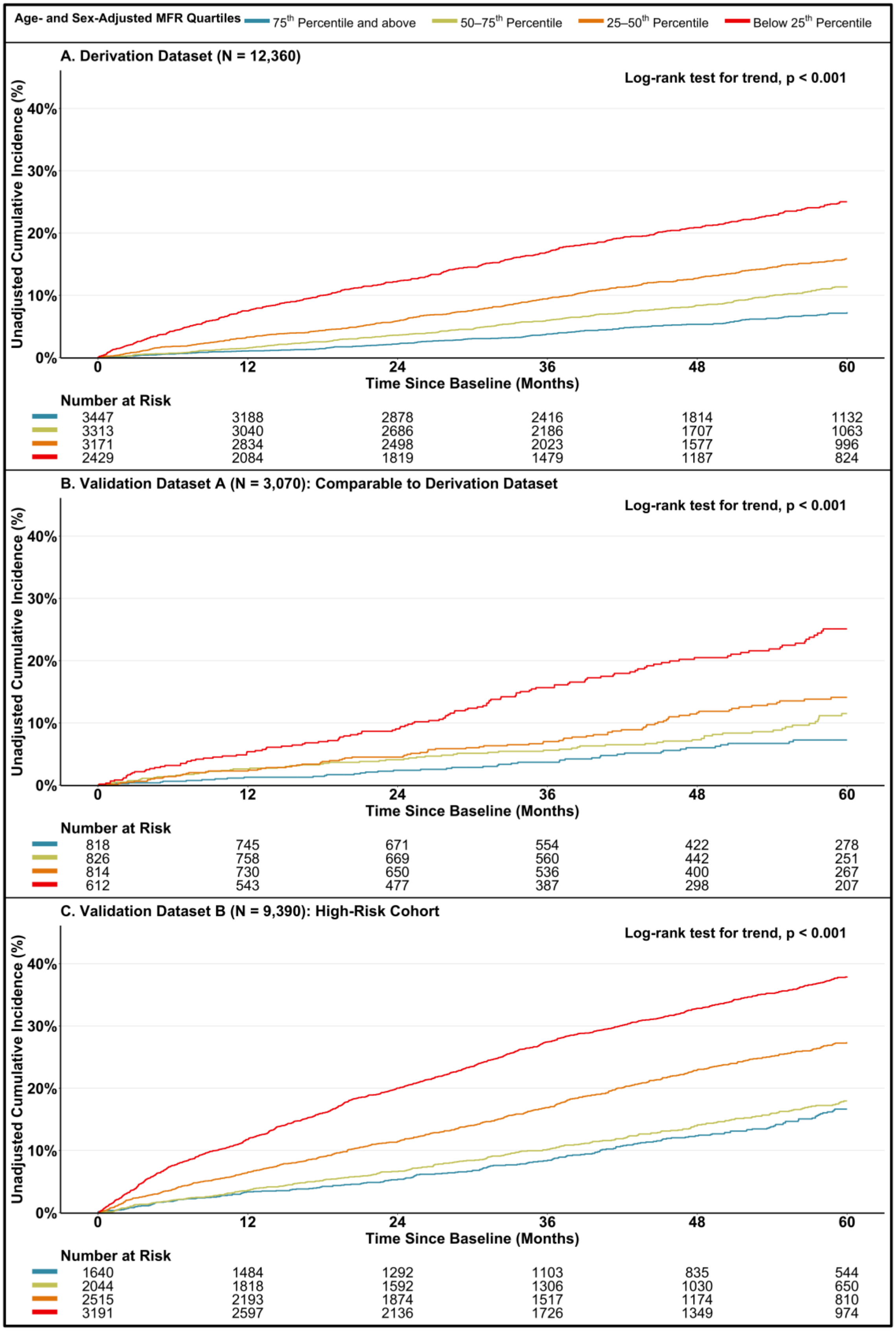

Supplemental Figure 8. Unadjusted Cumulative Incidence of a Composite of Cardiovascular (CV) Mortality, Hospitalization for Heart Failure (HF) or Myocardial Infarction (MI). Shown is the unadjusted 5-year cumulative incidence of a composite of CV mortality, HF or MI among individuals in the derivation (Panel A), validation A (Panel B) and validation B (Panel C) datasets according to quartiles of decreasing levels of age- and sex- adjusted MFR percentiles.

**Composite of CV Mortality, HF or MI**

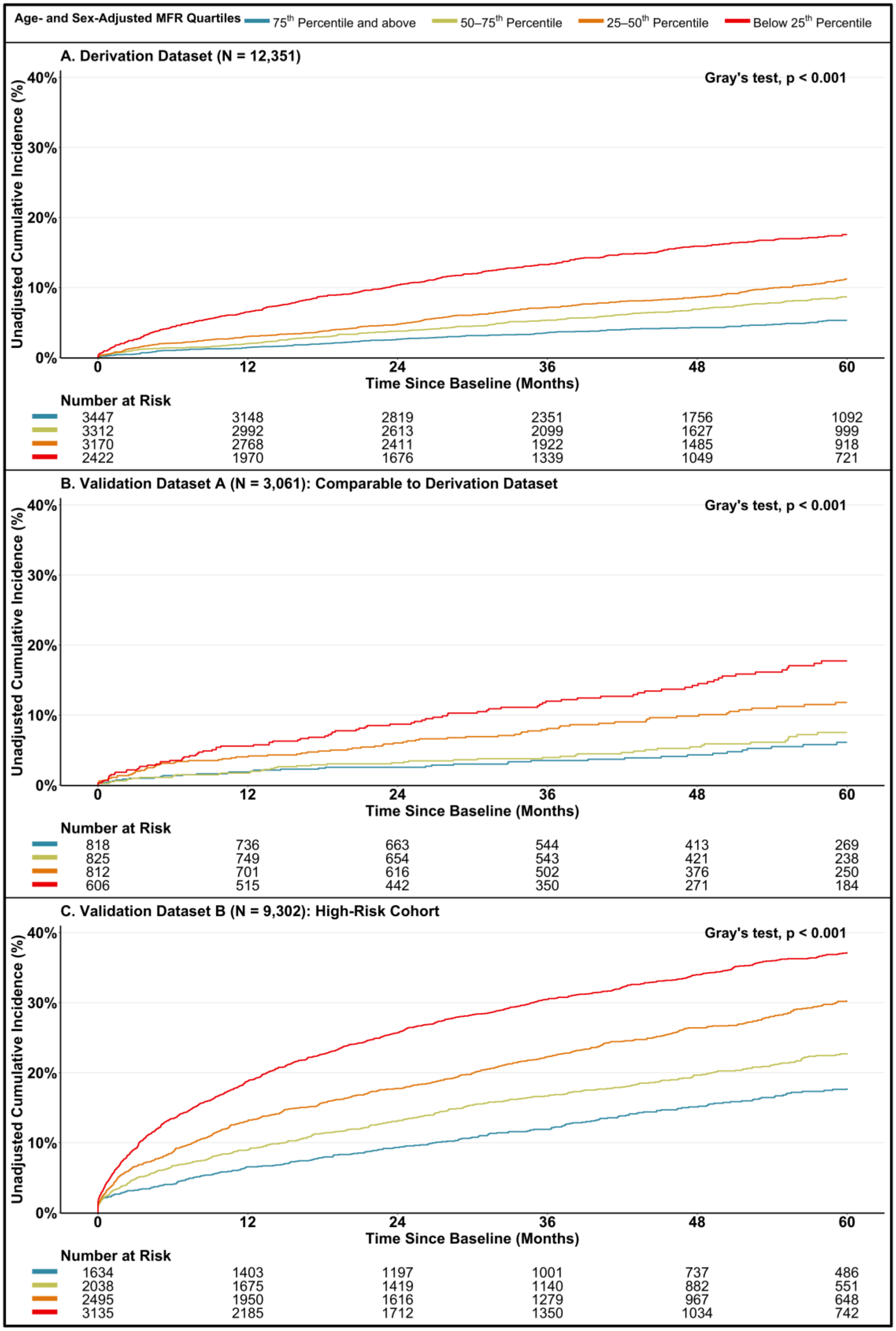

Supplemental Figure 9. Unadjusted Cumulative Incidence of Cardiovascular (CV) Mortality. Shown is the unadjusted 5-year cumulative incidence of a composite of CV mortality among individuals in the derivation (Panel A), validation A (Panel B) and validation B (Panel C) datasets according to quartiles of decreasing levels of age- and sex- adjusted MFR percentiles.

CV Mortality

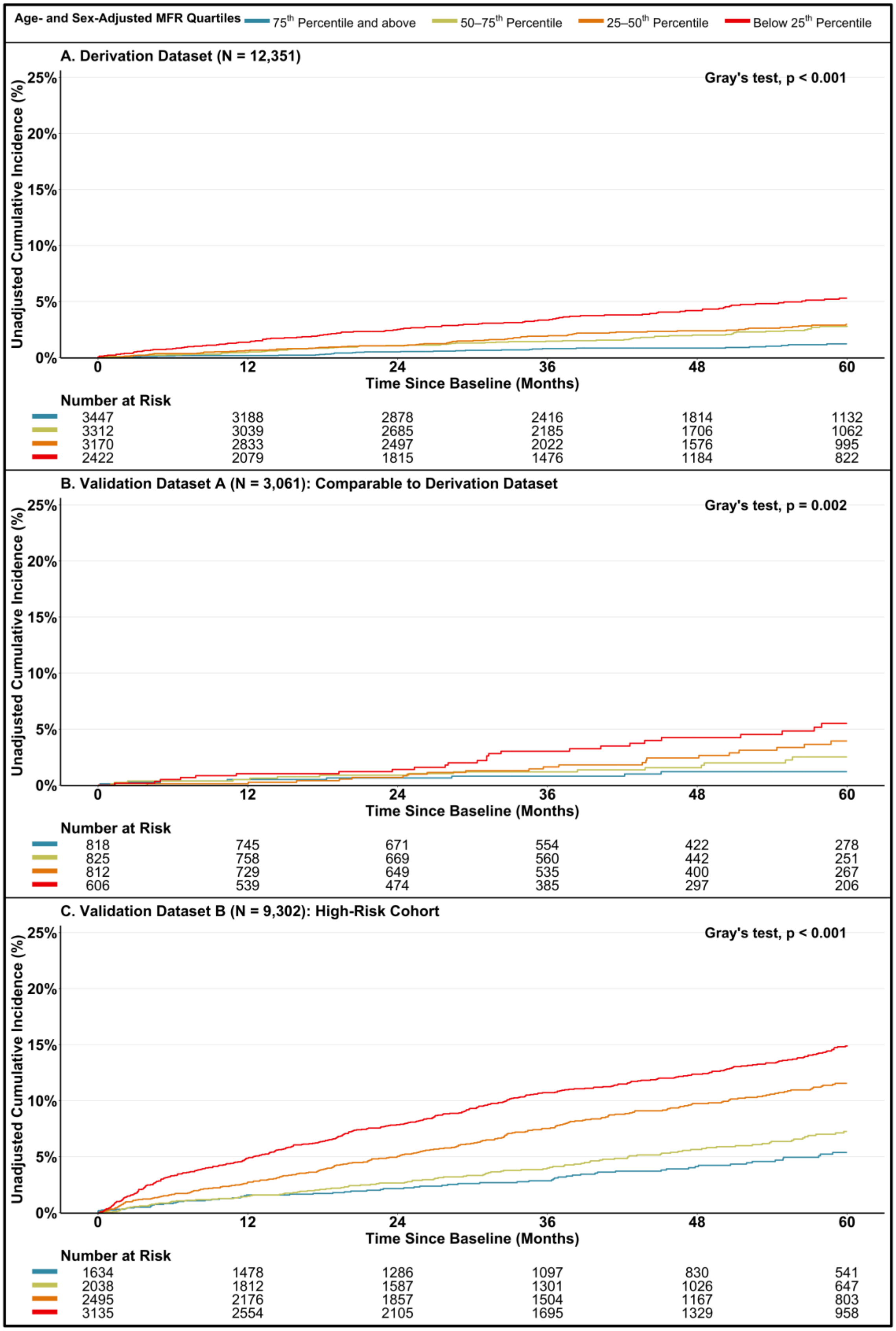
